# A systematic analysis of the contribution of genetics to multimorbidity and comparisons with primary care data

**DOI:** 10.1101/2024.05.13.24307009

**Authors:** Olivia Murrin, Ninon Mounier, Bethany Voller, Linus Tata, Carlos Gallego-Moll, Albert Roso-Llorach, Lucía A Carrasco-Ribelles, Chris Fox, Louise M Allan, Ruby M Woodward, Xiaoran Liang, Jose M Valderas, Sara M Khalid, Frank Dudbridge, Sally E Lamb, Mary Mancini, Leon Farmer, Kate Boddy, Jack Bowden, David Melzer, Timothy M Frayling, Jane AH Masoli, Luke C Pilling, Concepción Violán, João Delgado

## Abstract

**Background:** Multimorbidity, the presence of two or more conditions in one person, is common but studies are often limited to observational data and single datasets. We address this gap by integrating large-scale primary-care and genetic data from multiple studies to identify novel multimorbidity patterns and producing digital resources to support future research.

**Methods:** We defined chronic, common, and heritable conditions in individuals aged ≥65 years, using two large primary-care databases [CPRD (UK) N=2,425,014 and SIDIAP (Spain) N=1,053,640], and estimated heritability using the same definitions in UK Biobank (N=451,197). We used logistic regression to estimate the co-occurrence of pairs of conditions in the primary care data. Linkage disequilibrium score regression was used to estimate genetic similarity between pairs of conditions. Meta-analyses were conducted across databases, and up to three sources of genetic data, for each pair of conditions. We classified pairs of conditions as across or within-domain based on the international classification of disease.

**Findings:** We identified 72 chronic conditions, with 43·6% of 2546 pairs showing higher co-occurrence than chance in primary care and evidence of shared genetics. Many across-domain pairs exhibited substantial shared genetics (e.g. iron deficiency anaemia and peripheral arterial disease: genetic correlation *R_g_* =0·45[95% Confidence Intervals 0·27:0·64]). 33 pairs displayed negative genetic correlations, such as skin cancer and rheumatoid arthritis (*R_g_* =-0·14[-0·21:-0·06]), due to potential adverse drug effects. Discordance between genetic and primary care data was also observed, e.g., abdominal aortic aneurysm and bladder cancer co-occurred in primary care but were not genetically correlated (Odds-Ratio=2·23[2·09:2·37], *R_g_* =0·04[-0·20:0·28]) and schizophrenia and fibromyalgia were less likely to co-occur together in primary care but were positively genetically correlated (OR=0·84[0·75:0·94], *R_g_* =0·20[0·11:0·29]).

**Interpretation:** Most pairs of chronic conditions show evidence of shared genetics, and co-occurrence in primary care, suggesting shared mechanisms. The identified patterns of shared genetics, negative correlations and discordance between genetic and observational data provide a foundation for future multimorbidity research.

**Funding:** UK Medical Research Council [MR/W014548/1].

**Research in context:** *Evidence before this study:* We. Searched PUBMED, Embase, Global Health for articles published in English, since 6. Date >2012 (after the definitive Barnett et al. multimorbidity review paper was published) until 01 May 2024., using the search terms: (((multidisease*[tiab] OR multi-disease*[tiab] OR 3ultimorbidity*[tiab] OR multi-morbidit*[tiab] OR multipatholog*[tiab] OR multi-patholog*[tiab] OR pluripatholog*[tiab] OR polypatholog*[tiab] OR poly-pathology*[tiab] OR “multiple long-term conditions”[tiab])) OR (multimorbidity[MeSH Terms])) AND (“genetic*”[tiab] OR “genomic*”[tiab] OR (“genome wide association stud*”[tiab] OR “genome wide association analysis”[tiab] OR “GWAS”[tiab])) NOT (animals[mh] NOT humans[mh]) AND NOT (“retracted publication”[pt]). We excluded studies investigating monogenic mutations and case studies. 243 studies met the primary inclusion criteria, where 68 presented original research addressing multimorbidity and incorporating genetic analysis. Multimorbidity is predominantly characterised by counts or clusters of conditions, with limited exploration of underlying mechanisms. Only 11 investigated multimorbidity beyond the analysis of comorbidities of single conditions, with five reporting systematic investigations of genetics across multiple long-term conditions. None of these studies compare data from more than one genetic database with ‘real-world’ data. This comparison is necessary to identify correlations of diseases and to be able to carry out preventive actions that facilitate better health care.

*Added value of this study:* We describe a large-scale analyses into the genetics of multimorbidity informed by representative primary care data, starting from a systematic standardisation of relevant diagnostic codes, and the ascertainment of conditions harmonised across multiple databases. Importantly, we include the analysis of multiple sources of data from both observational and genetic databases, whereas previous studies have usually been limited to one study, such as UK Biobank. We also produced a detailed interactive interface for 72 common long-term and heritable conditions and 2546 pairs of conditions across multiple sources of data: https://gemini-multimorbidity.shinyapps.io/atlas. Together we describe the parallel analysis of specific co-occurring pairs in primary care and genetic data, involving 100,000s to millions of patients and 2546 pairs of conditions. These comparisons identified novel pairs of conditions, and pairs of conditions that shared genetics but did not co-occur in primary care, or vice versa.

*Implications of available evidence:* Large-scale, digital databases of electronic medical records linked to genetic information involving 100,000s of patients can significantly enhance our comprehension of multiple long-term conditions (multimorbidity). This advancement builds on previous work by providing evidence of potential shared biology between 100s of specific pairs of conditions.

## Introduction

Multimorbidity is the coexistence of two or more long-term conditions (LTCs) and is a growing problem globally, projected to affect two-thirds of 65-year-olds in the next decade.^1^ Multimorbidity is a clinical marker for accelerated ageing, frailty and mortality. Treatment of multiple co-existing LTCs is a growing, disproportionate economic and capacity burden for healthcare systems.^2,3^ The mechanisms driving the accumulation of LTCs remain uncertain, and despite recent efforts clinical research to understand, prevent and treat multimorbidity is needed.

Most research into multimorbidity is observational, focusing on the concurrence of LTCs as raw or weighted counts of selected conditions. Fewer studies used clustering approaches to reveal LTC groupings (e.g., cardiovascular and metabolic) with variability in included conditions and identified clusters.^4^ Clustering-based approaches, favour high-prevalence LTCs (e.g., hypertension) while low prevalent LTCs, and LTC pairs being underrepresented in analyses.^5,6^ Alternative approaches focussed on pairs of conditions, which are the constituents of multimorbidity, and analysed disease pairs using genetics to identify potential shared pathways;^7–9^ however, focusing on small sets of closely related conditions, single large datasets such as the UK Biobank,^7^ or genetic correlations across multiple, broadly-defined traits without direct comparison to real-world primary-care datasets.^8,9^

Genetic data provide opportunities to understand underlying mechanisms with less susceptibility to confounding, biases, and reverse causality than observational studies. Summary-level genetic association data is available for many common LTCs based on studies including 10,000s to 100,000s cases. These data enable the comparison of LTCs across, as well as within, patients, and datasets. For example, linkage disequilibrium (LD) score regression is an approach that can be used to compare genetic similarity between conditions from separate case-control studies of separate diseases, even when each study has no information on the other condition.^10,11^ A systematic, multi-modal data investigation remains necessary to identify potential biological mechanisms and opportunities for the prevention and treatment of multimorbidity.

In this study, we combine the advantages of healthcare datasets (representativeness and large sample sizes) with the advantages of genetic studies (large sample sizes, less confounding or reverse causality) to study multimorbidity. In contrast to most previous studies, we analyse specific pairs of LTCs, rather than counts or clusters of conditions, and meta-analyse data from multiple sources. We provide a comprehensive assessment of the genetic and observational relationships between 72 LTCs common in people aged over 65 years - an analysis comprising 2,546 pairs. Our study identifies hundreds of pairs of conditions with shared genetic factors, many from across traditional disease domains; compares genetic evidence of the co-occurrence of conditions with observational evidence from healthcare data; and describes a powerful new resource for the study of multiple long-term conditions in multiple databases.

## Methods

### Design and populations Primary-care healthcare data

Observational data were obtained from two independent databases of electronic health records: 1) The UK Clinical Practice Research Datalink (CPRD), including data from >30 million National Health Service (NHS) patients from >700 primary-care practices.^12^ 2) The Spanish Information System for Research in Primary Care (SIDIAP), including data from 6 million patients from 328 practices.^13^ We included N=2,425,014 (CPRD) and N=2,425,014 (SIDIAP) patients alive, registered and over 65 years of age, and age group associated increase prevalence for LTCs. Clinical records in both the UK and Spain undergo standardised coding to contribute to disease registers and healthcare planning. In SIDIAP the electronic codes are recorded using the International Classification of Disease, version 10 (ICD-10). In CPRD codes are a mixture of Read v2 Codes, EMIS and SNOMED codes.

### Genetic

Genetic data from 3 sources were included 1) UK Biobank (UKB),^14^ a large cohort study with 500,000 individuals with baseline data linked to electronic health records; we used 450,197 individuals genetically similar to the 1000 Genomes European reference population (“EUR-like” - see Supplementary Information). 2) FinnGen, a large-scale genomics initiative linking diagnosis to genotype data in 377,277 participants (release 9).^15^ 3) Published disease-specific Genome Wide Association Study (GWAS) meta-analyses summary statistics. For these disease-specific GWAS, we used the European Bioinformatics Institute (EBI) GWAS Catalog,^16^ disease-specific public repositories, and, if necessary, contacted authors of GWAS to obtain summary association statistics.

### Defining Long-Term Conditions

LTCs were defined and selected for analysis based on chronicity, prevalence, and heritability as detailed in the Supplementary Methods, and summarized below. Selected conditions were assigned to disease domains based on the ICD-10 chapters: https://icd.who.int/browse10/2019/en.

#### Step 1: Selection of conditions – chronicity

We included only LTC or with sequelae lasting more than 3 months to meet the definition of multimorbidity. The definition of disease chronicity was adapted from chronic conditions published by Calderón-Larrañaga et al.^17^ The LTC code lists were compared with CALIBER interoperable code lists,^18^ and adapted with clinician input to refine and remove acute diagnostic codes. This process generated 232 LTC code lists (Supplementary Figure 1).

#### Step 2: Selection of conditions – prevalence

Prevalence was estimated using CPRD and SIDIAP data for individuals aged 65 and older who were alive and registered on the 1^st^ of January 2020. A LTC was diagnosed if a diagnosis code was recorded at any time before 1st January 2020. These databases are population-representative sources of primary-care data, providing the most comprehensive set of long-term conditions, given many are not treated in hospital. Secondary care data was excluded as it is not consistently available in SIDIAP. The cutoff date minimises the impact of the COVID-19 pandemic in our analyses. We included conditions with a prevalence greater than 0.5%, supplemented by clinician and Patient and Public Involvement and Engagement (PPIE) resulting in 84 LTCs out of 232 proceeding to step 3 (see PPIE section below).

#### Step 3: Selection of conditions - Heritability

We performed GWAS for 84 LTCs in UKB, using our defined clinical code lists and the REGENIE software (v3.1.3).^19^ See the Supplementary Information for details. Briefly, analyses were adjusted for age, sex, genotyping chip, and assessment centre. We restricted genetic variants to those with a minor allele frequency (MAF) of >0·1%, and an imputation INFO score ≥0·3. SNP-based heritability was estimated using GWAS summary statistics and LD-score regression (LDSC).^10^ We used the 1000 Genomes EUR reference population LD data throughout. 72 LTCs met criteria for analysis (Supplementary Figure 1).

### Statistics

#### Co-occurrence of LTCs in primary-care

Logistic regression models tested the likelihood of two LTCs co-occurring in observational data, with models adjusted for age and sex, a Benjamini-Hochberg correction was used to account for multiple testing (additional detail in supplemental information). Associations (i.e. odds-ratios) between LTCs were estimated in CPRD AURUM and SIDIAP, and meta-analysed with fixed-effects – the appropriate model for few data sources (N=2).^20^Q tests estimated heterogeneity in the association between databases (Supplementary Table 4). Results stratified by country are available online https://gemini-multimorbidity.shinyapps.io/atlas.

#### Genetic Correlation

To provide the most powerful set of genetic data we sought to meta-analyse genetic studies according to two criteria: first, there should be evidence that the conditions defined in the different genetic studies were the same, or very similar, conditions. Second, there should be no overlap between the sources of genetic data. We used LDSC to estimate the within-condition between-dataset genetic correlation (R) and limited meta-analyses to studies where the within-condition genetic correlation was >0·8.^11^ To ensure there was no overlap in genetic data, The FinnGen and Consortium data were added to the meta-analysis when within-condition R with UKB was >0·8, otherwise we used UKB only (i.e., the genetic evidence suggests that the conditions used by consortium GWAS and/or FinnGen are not consistent with those defined using diagnostic codes for LTCs in our project for UK Biobank). Where Consortium data included UKB and/or FinnGen, Consortium data were favoured, because of the larger number of cases, and UKB/FinnGen excluded to avoid sample overlap. Studies were meta-analysed using GWAMA.^21^ LTC pairs with genetic correlation at a false discovery rate of 5% were first separated into those within and across domains and second, sorted into three groups as those with weak, intermediate, and strong genetic correlations. See the Supplementary Methods for individual study references, Supplementary Figure 2 for analysis flowchart, and Supplementary Table 1 for effective sample size and other information.

#### Comparison between observational and genetic correlations

Linear regression models estimated the association between genetic correlations and observed co-occurrence between LTC pairs, where LTC pair genetic correlation was used as the dependent variable and observed co-occurrence as the outcome. Additional models estimated the associations in subsets of LTC pairs within and across disease domains. LTC pairs were classified as within or across disease domains based on ICD-10 chapters (Supplementary Table 3).

#### Colocalised gene analysis

For selected LTC pairs (see Results section) we: 1) defined genetic regions with variants significantly associated with both LTCs (i.e., lead SNPs p<5*10-8 separated by <250kbp); 2) used R package ‘coloc’^22^ (v5.2.3) to estimate the posterior probability (using ‘coloc’ default priors) that the pair of LTCs share a single causal variant in the region (“PP4”); 3) where there was high evidence for a shared causal region (PP4>0.9) we investigated the likely shared mechanism using the individual candidate variants highlighted by coloc. We identified the likely causal gene (the nearest), known disease associations (DisGeNET & GWAS-catalog),^16,23^ and whether it is a known or potential drug target (using DrugBank and other resources).^24^

#### Professional, Patient and Public Involvement

Two co-authors (LF and MM) are public collaborators with direct experience of living with multiple LTCs. They are co-investigators attending fortnightly research meetings to co-develop the research. Additional workshops for patients and carers with experience of multiple LTCs have contextualised the importance of the research and directly informed research decisions on LTC selection and refinement as outlined above.^25^

Healthcare professionals, including primary and secondary care physicians and allied healthcare professionals (led by authors JM, CVF and SEL) informed the definition, selection, and precision coding of LTCs. Detailed clinician review of LTC pairs identified potential underlying mechanisms and selected LTC pairs where an association was considered novel.

#### Ethics

This study was approved by the relevant ethics committees: SIDIAP Scientific and Ethical Committees (19/518-P) on 18/12/2019. The SIDIAP database is based on opt-out presumed consent. If a patient decides to opt out, their routine data would be excluded of the database.

#### CPRD ISAC committee protocol number 23_003109

The Northwest Multi-Centre Research Ethics Committee approved the collection and use of UK Biobank data for health-related research (Research Ethics Committee reference 11/NW/0382). UKB was granted under Application Number 9072.

## Results

### The majority of long-term conditions common in older people are heritable

We identified 72 LTCs that were common and showed evidence of heritability. The most prevalent of these conditions in primary-care data were hypertension (CPRD: 51·7%; SIDIAP: 63·2%), osteoarthritis (CPRD: 35·3%; SIDIAP: 37·8%), and upper body enthesopathy (CPRD: 27·0%; SIDIAP: 20·0%) (Supplementary Table 1). The most heritable of these LTCs were fibromatoses (h^2^=25·1%) and type 2 diabetes (h^2^=21·3%) (Supplementary Table 1). Further details of all 72 plus 12 additional conditions that did not meet our heritability criteria are available online: https://gemini-multimorbidity.shinyapps.io/atlas.

### Associations between conditions

Pairwise combinations of 72 conditions resulted in 2,546 pairs, of which 260 were within-domain and 2,286 across-domain (based on ICD-10 classification) (Figure 1; Table 1). Ten pairs were excluded due to overlapping code lists, e.g., transient ischaemic attack (TIA) and “all stroke”. Prevalence estimates were based on data from 2,425,014 CPRD patients (46·3% male) and 1,053,640 SIDIAP patients (43·0% male). Genetic correlations were estimated using data from 451,240 UKB participants (45·7% male), 377,277 FinnGen participants (44·1% male), and from condition-specific GWAS data with sample sizes ranging from 12,366 to 181,522 cases (Supplementary Table 1-2).

**Figure 1.**
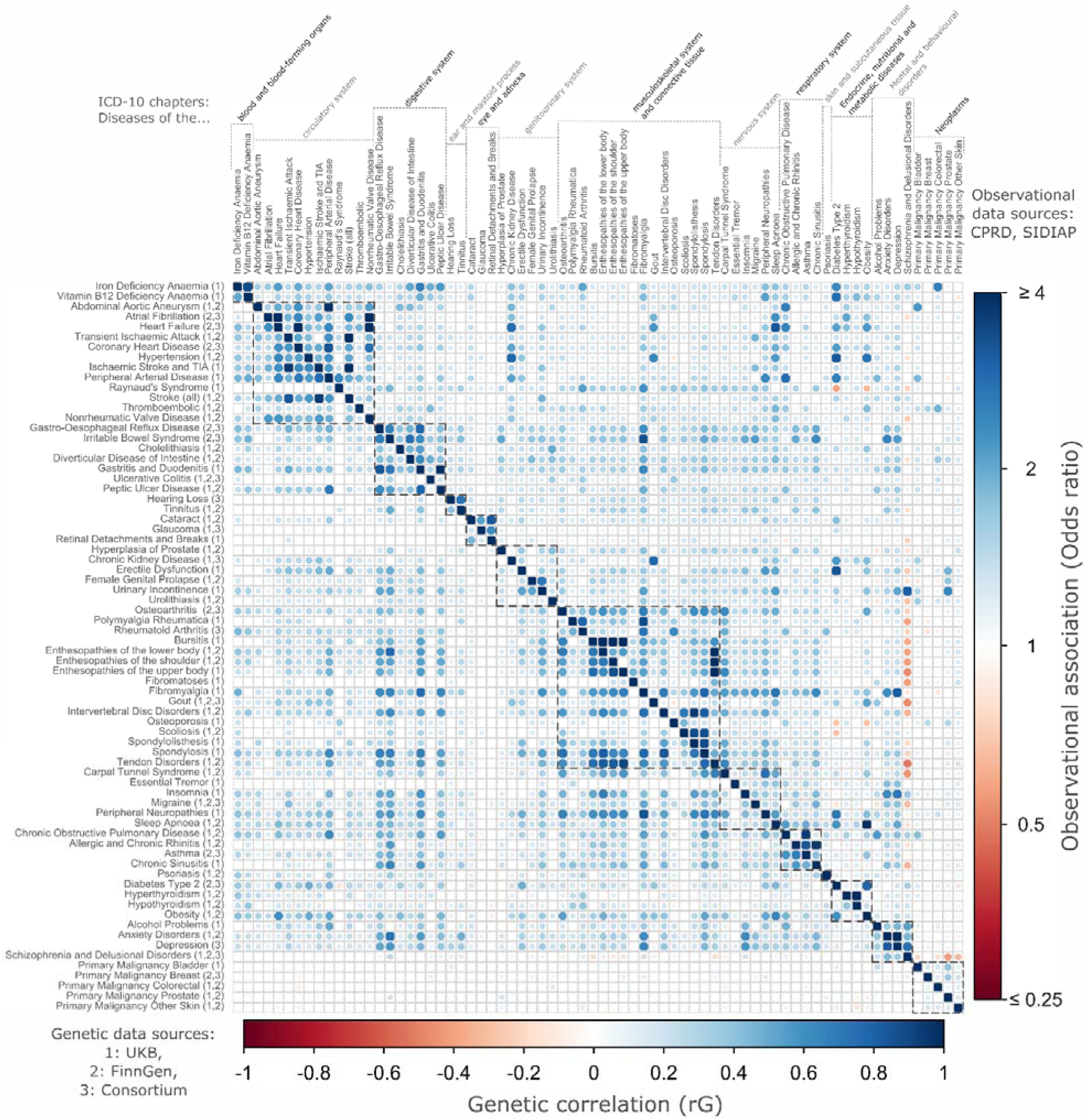
Pairwise associations between 72 long-term chronic conditions: co-occurrence in observational data (upper right panel) and genetic correlation (lower left panel) LTCs tested are common (≥0.5% prevalence in individuals aged 65 and older) and heritable (SNP-based heritability z-score >4) – see Methods. Observational associations (upper right triangle of heatmap) are Odds Ratios from meta-analysed logistic regression models from two population-representative primary care cohorts (CPRD and SIDIAP) comprised of individuals aged 65 or older and alive on January 1^st^, 2020 (to aid in readability, ORs >4 are set to 4, and ORs <0·25 are set to 0·25). Genetic correlations (lower right triangle of heatmap) are from meta-analysed GWAS summary statistics from up to three sources (UKB, FinnGen, and published consortium studies). LTCs within the same ICD-10 chapter (i.e., “within-domain”) are highlighted with dotted lines. See Supplementary Tables 4 and 5 for full results, and web app for interactive version: https://gemini-multimorbidity.shinyapps.io/atlas.

**Table 1.** Observational co-occurrence and genetic correlation between LTC pairs.

| All | Observational (primary care) associations |  |  |  |  |
| --- | --- | --- | --- | --- | --- |
| Genetic Correlation | Not Significant | Weak | Intermediate | Strong | Total |
| Not Significant | 100 (10.8) | 765 (82.7) | 53 (5.7) | 7 (0.8) | 925 (36.3) |
| Weak | 18 (2.1) | 692 (79.8) | 138 (15.9) | 19 (2.2) | 867 (34.1) |
| Intermediate | 0 (0) | 265 (45.8) | 245 (42.3) | 69 (11.9) | 579 (22.7) |
| Strong | 0 (0) | 18 (10.3) | 69 (39.4) | 88 (50.3) | 175 (6.9) |
| Obs Total | 118 (4.6) | 1740 (68.4) | 505 (19.8) | 183 (7.2) | 2546 (100) |
| Across-domain Pairs | Not Significant | Weak | Intermediate | Strong | Total |
| Not Significant | 98(4.3) | 727(31.8) | 45(2) | 6(0.3) | 876 (38.3) |
| Weak | 16(0.7) | 653(28.6) | 115(5) | 12(0.5) | 796 (34.8) |
| Intermediate | 0 (0) | 256(11.2) | 209(9.1) | 44(1.9) | 509 (22.3) |
| Strong | 0 (0) | 18(0.8) | 51(2.2) | 36(1.6) | 105 (4.6) |
| Obs Total | 114 (4.9) | 1654 (72.4) | 420 (18.4) | 98 (4.3) | 2286 (100) |
| Within-domain pairs | Not Significant | Weak | Intermediate | Strong | Total |
| Not Significant | 2(0.8) | 38(14.6) | 8(3.1) | 1(0.4) | 49 (18.9) |
| Weak | 2(0.8) | 39(15.0) | 23(8.8) | 7(2.7) | 71 (27.3) |
| Intermediate | 0 (0) | 9(3.5) | 36(13.8) | 25(9.6) | 70 (26.9) |
| Strong | 0 (0) | 0 | 18(6.9) | 52(20) | 70 (26.9) |
| Obs Total | 4 (1.5) | 86 (33.1) | 85(32.7) | 85(32.7) | 260 (100) |

For each pair the log-odds ratio of the least prevalent disease explained by the more prevalent disease adjusting for age and sex was meta-analysed across CPRD and SIDIAP. Observed LTC pairs were divided into terciles based on within-domain pairs; weak (Odds-Ratio [OR] $\leq 1.46$ ) intermediate ( $1.46 < \text{OR} \leq 1.90$ ) and strong ( $\text{OR} > 1.90$ ). The genetic correlations of LTC pairs were divided into terciles based on within-domain pairs: weak ( $R_g \leq 0.25$ ), intermediate ( $0.25 < R_g \leq 0.48$ ) and strong ( $R_g > 0.48$ ) correlation.

Regression analyses across N=2,546 pairs demonstrated that a 1% increase in R equates to a 2·76% increase in the odds of co-occurrence (95% Confidence Intervals 2·65%-2·87%), thus the stronger the genetic correlation between an LTC pair, the higher the chance of the pair co-occurring in primary-care.

## 1. The majority of within-domain LTC pairs are genetically correlated and co-occur together in primary-care data

From 260 pairs of within-domain LTCs, N=209 (80·4%) were genetically correlated and co-occurred in primary-care more often than expected (e.g., heart failure and atrial fibrillation - *R_g_* =0·60-SE=0·03, OR: 7·53 [7·45-7·61]). Significant negative genetic correlations were absent in this group (Figure 2, blue).

**Figure 2.**
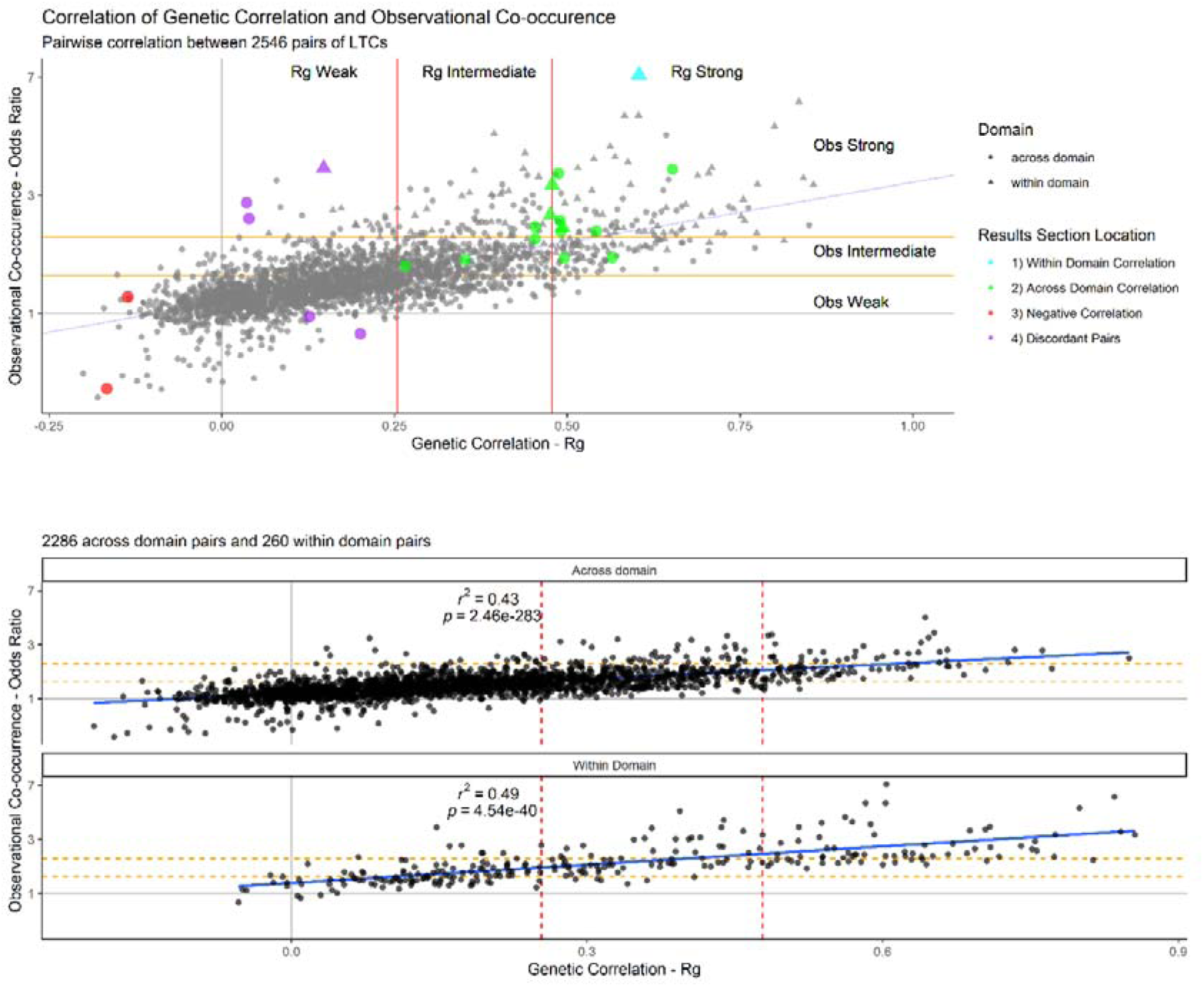
The relationship between LTC observational co-occurrence and genetic correlation Scatter plots of the relationship between observed co-occurrence and genetic correlation of LTC pairs. (blue) linear regression line, (yellow) terciles of genetic correlation, (red) terciles of likelihood of observed co-occurrence. Terciles estimated based on within-domain LTC pairs. The upper panel shows all pairs with pairs discussed in result section 1:4 highlighted. The lower two panels show the across-domain (left, n=2,286) and within-domain (right, n=260) pairs. See Supplementary Tables 4 and 5 for full results, Supplementary Table 6 for the highlighted pairs and the web app for interactive versions: https://gemini-multimorbidity.shinyapps.io/atlas. See Supplementary Figure 3 for plot stratified by domain.

## 2. Many across-domain LTC pairs are genetically correlated and tend to co-occur in primary-care data

In across-domain pairs, we identified N=105 (4·6%) that were as strongly genetically correlated R >0·48) as the strongest third of within-domain pairs. These pairs were more likely to co-occur in primary-care (Figure 2, green). For example, sinusitis and gastro-oesophageal reflux disease (GORD) *R_g_* =0·49-SE=0·06), erectile dysfunction and peripheral neuropathy (*R_g_* =0·49-SE=0·10) and iron deficiency anaemia and peripheral arterial disease (*R_g_* =0·45-SE=0·10) were as strongly correlated as within-domain pairs coronary heart disease and stroke (*R_g_* =0·49-SE=0·03), rheumatoid arthritis and polymyalgia rheumatica (*R_g_* =0·48-SE=0·09), and TIA and peripheral arterial disease (*R_g_* =0·48-SE=0·08). Diseases of the musculoskeletal system or connective tissue were present in 8 of the 10 across-domain LTC pairs with the strongest genetic correlations, but only 18·3% of all significant across-domain pairs.

Clinician reviews showed that all within-domain LTC pairs with shared genetics are known to clinicians and have widely accepted explanations. Clinician reviews of across-domain pairs with shared genetics showed that the majority have accepted explanations. For example, associations could reflect 1) shared pathology (e.g. type 2 diabetes and erectile dysfunction – OR: 3·27 [3·24-3·30], *R_g_* =0·49-SE=0·05), 2) the first condition is a risk factor for the second (e.g. obesity and osteoarthritis – OR: 2·00 [1·99-2·01], *R_g_* =0·54-SE=0·02), 3) treatment effects (e.g. intervertebral disc herniation where non-steroidal anti-inflammatory drugs (NSAIDs) could pre-dispose to GORD - OR: 1·60 [1·58-1·62], *R_g_* =0·50-SE=0·03) and 4) shared presenting symptoms leading to overlapping diagnoses (e.g., sinusitis and GORD - OR: 2·00 [1·98-2·02], *R_g_* =0·49-SE=0·06). However, other across-domain LTC pairs had no well-established shared mechanisms for genetic and observed associations.

Some examples were tendon disorders and diverticular disease (OR: 1·49 [1·48-1·50], *R_g_* =0·27-SE=0·03), fibromyalgia and irritable bowel syndrome (IBS) (OR: 3·38 [3·30-3·47], *R_g_* =0·65-SE=0·13), fibromyalgia and asthma (OR: 1·87 [1·83-1·92], *R_g_* =0·45-SE=0·06) and IBS and peripheral neuropathy (OR: 1·60 [1·57-1·64], *R_g_* =0·57-SE=0·12). A total of N=89 genetically correlated and co-occurring LTC pairs involved treatable deficiencies in iron and vitamin B12, e.g. B12 deficiency and COPD (OR: 1·57 [1·55-1·60], *R_g_* =0·35-SE=0·06).

## 3. A small number of pairs of conditions are negatively genetically correlated and co-occur less often than expected in primary-care

We identified 33 (1·3% of N=2,546) pairs of LTC conditions that were negatively genetically correlated (Q-value <0·05), implying that the genetic risk of one is associated with protection from the other. Nineteen (56·0%) of these pairs were observed together less often than expected in primary-care. Of the 19 pairs, 16 involved malignancies, for example, skin cancer and rheumatoid arthritis (*R_g_* =-0·14-SE=0·04) and five pairs included schizophrenia (e.g. schizophrenia and upper body enthesopathy, *R_g_* =-0·17-SE=0·03) (Figure 2, red).

## 4. Some pairs of conditions show discordance between observed co-occurrence in primary care and genetic correlation

A total of 34 (1·3% of 2,546) pairs did not co-occur in primary-care data more than chance (after false discovery rate correction) but had a positive genetic correlation (Figure 2, purple), such as female genital p6rolapse and type 2 diabetes (OR: 0·97 [0·96-0·98], *R_g_* =0·13-SE=0·02). This group included several pairs involving mental health conditions and pain-related conditions e.g. schizophrenia and fibromyalgia (OR: 0·84 [0·75-0·94], *R_g_* =0·20-SE=0·04).

We identified 739 disease pairs (29·0%) showing significant positive co-occurrence in primary care but with non-significant genetic correlation (Q-value ≥0·05) (Figure 2, purple). Examples include fibromyalgia and polymyalgia rheumatica, indicative of explorative diagnosis along a diagnostic pathway (OR: 3·41 [3·28-3·54], *R_g_* =0·15-SE=0·12-Q=0·30); iron deficiency anaemia with colorectal cancer (OR: 2·55[2·49-2·60], *R_g_* = 0·04-SE=0·07-Q=0.66), as well as abdominal aortic aneurism (AAA) with bladder cancer (OR: 2·23[2·09-2·37], *R_g_* =0·04-SE=0·12-Q=0·80), which may involve incidental findings.

## 5. Mechanisms highlighted by genetic colocalisation for specific LTC pairs

To illustrate the approach to investigating the mechanism linking LTC pairs that are genetically and observationally correlated in our analysis, we selected one “within domain” pair (atrial fibrillation [AF] and heart failure [HF], as a positive control) and all 17 “across domain” pairs from Supplementary Table 6. We found high evidence (PP4>0.9) for colocalisation at 3 regions for AF and HF; though all are known AF loci, most published AF loci did not have high probability of colocalising with HF. Therefore, these are promising for follow up, for example CAMKD2, which has previously been studied as a target for Rheumatoid arthritis (https://go.drugbank.com/bio_entities/BE0004108). In the “across domain” pairs, we found high evidence for a colocalising region between 3 pairs (summarized below), and include further details in Supplementary Table 6.

### 5.1 Osteoarthritis and obesity

We estimated the role of obesity as a shared modifiable risk factor between LTC pairs in our parallel study^26^, demonstrating that for some LTC pairs (e.g., type-2 diabetes [T2D] and OA) BMI fully attenuated the genetic correlation.

### 5.2. Rheumatoid arthritis (RA) and non-melanoma skin cancer

One variant (rs3087243) had high evidence of colocalisation (PP4=1) and is in close proximity (<200bp) from gene CTLA4. CTLA4 is a target for FDA-approved drugs used to treat several cancers (e.g., ipilimumab) and Rheumatoid arthritis (abatacept). Observationally, RA and skin cancer were positively correlated (OR 1.15, p=4*10^-23^) however, in our genetic analysis we observed a negative genetic correlation (rG −0.14, p=5*10-4). Consistent with this, we found that the rs3087243 G-allele increased risk of RA (per allele OR 1.09, p=3*10^-19^) but decreased risk of skin cancer (per allele OR 0.94, p=4*10^-17^).

### 5.3. T2D and Female Genital Prolapse (FGP)

One region had high evidence of colocalisation (PP4=1), with 5 possible variants (rs2267373 had highest likelihood: PPH4=0.22): these are intronic for genes MAFF and PLA2G6, and have previously been identified in GWAS of cholesterol and related traits (REF).^16^ Neither protein is a known drug target in DrugBank, though PLA2G6 is a phospholipase, again highlighting the links with cholesterol. The genetic correlation was independent of the causal effect of BMI in our parallel analysis. ^26^

## Discussion

### Associations between conditions and mechanisms of multimorbidity

We report a large, systematic investigation of the shared genetics and co-occurrence in primary-care of 2546 pairs of long-term conditions (LTC). Our study builds on previous work in multimorbidity by integrating two of the largest most representative sources of primary-care data with large-scale genetic data for each of 72 conditions. Most pairs (61·0%) tended to co-occur in primary-care data and share genetics, with a subset of across-domain pairs showing genetic correlations as strong as many within-domain pairs. The overall positive relationship between observed phenotypic associations and genotypic correlations is consistent with and considerably expands work from a previous smaller-scale study, limited to 17 conditions in UK Biobank.^27^ These findings suggest diverse shared pathways and mechanisms drive co-occurrence of LTCs in multimorbidity.

There are several potential mechanistic explanations for our results. These mechanisms could include shared pathophysiology, such as atherosclerosis, as a likely shared risk factor for cardiovascular diseases such as atrial fibrillation and heart failure. Causal mechanisms may also include one LTC acting as a risk factor for a second LTC, such as obesity increasing the risk of osteoarthritis due to increased mechanical stress on weight-bearing joints. We extended this concept further with evidence that the genetics of BMI fully explains the shared genetics between 7% of studied LTC pairs, such as T2D and OA, and partially for 30% (e.g., hypertension and sleep apnoea *R_g_* = 0·40 reduces to *R_g_* = 0·30 after removing the causal effect of BMI on both conditions, difference-p=9*10^-24^).^26^ Shared genetic and observational mechanisms between LTCs could result from the combined effect of both types of causal pathways, such as increased risk of erectile dysfunction in type 2 diabetes, caused by shared pathophysiology and by the effect of hyperglycaemia on the endothelium.^28^ Lastly, concordance between phenotypic and genetic correlations may result from iatrogenic mechanisms. For example, secondary stroke prevention with antiplatelet medications, such as clopidogrel, increases the risk of gastritis, and the use of non-steroidal anti-inflammatory drugs (NSAIDs) for pain in musculoskeletal conditions such as intervertebral disc herniation increases the risk of gastro-oesophageal reflux disease (GORD).^29^

We highlight LTC pairs that co-occur more often than expected by chance and that are genetically correlated but lack strong clinical understanding. For example, tendon disorder and diverticular disease were associated and novel. A few LTC pairs include a readily treatable condition such as B12 deficiency or iron deficiency anaemia (IDA), highlighting a potential direct path towards intervention.

LTC pairs showing discordance between genetic correlations and co-occurrence in primary care raise interesting questions, including about clinical service provision. The 34 LTC pairs with evidence of shared genetics but occurring less often than expected by chance were dominated by combinations of musculoskeletal and mental health conditions, such as schizophrenia with fibromyalgia, and with rheumatoid arthritis, or alcohol addiction and spondylolisthesis. These combinations could suggest that diagnoses of severe mental health conditions lead to underdiagnosis of concomitant physical LTCs probably involving diagnostic overshadowing.^30^ LTC pairs with increased observed co-occurrence but without evidence of genetic correlation could include clinical instances where diagnosis with one LTC leads to increased odds of being diagnosed with a second LTC. For example, IDA and colorectal cancer, where a diagnosis of IDA triggers an investigation for colorectal cancer; or Abdominal Aortic Aneurysm (AAA) and bladder cancer, where an AAA is incidentally detected during an investigation for bladder cancer. Discordant pairs highlight instances where pathways for diagnosis of chronic conditions are inadequate, and these may remain undiagnosed and untreated.

### Strengths and limitations

This study uses large and representative data from electronic health records (EHRs), with findings replicated across datasets.^12,13^ EHRs are inclusive of those with disability, frailty and MLTC who are frequently under-represented in clinical research. Observational data were also complemented with three high-quality genetics data sources. An exhaustive review process has been carried out to compare chronic, heritable diseases across genetic and EHR data. There is no clear international consensus on the assessment of multimorbidity. In this study, we have used a comprehensive method to select common chronic LTCs in older populations, based on existing literature, that has ensured the inclusion of 2546 multimorbidity disease pairs, the largest systematic collection to date. Clinical experts curated LTC definitions and LTC pairs, which are available for future MLTC investigations.

A few limitations should be considered. 1) The genetic analyses were limited to individuals of European genetic ancestry due to a paucity of large-scale genetic data from people of non-European ancestry. 2) We presented a descriptive analysis of the correlation between genetic similarity and observed co-occurrence. The causal mechanisms for these relationships vary across LTC pairs, and additional work is required to investigate them. 3) Limiting primary-care data to individuals at least 65 years may introduce survival bias; negative effect sizes cannot be automatically conferred as protective. For example, where we found reduced co-occurrence between schizophrenia and musculoskeletal condition another study found an increased rate of mortality between the two.^31^ However, the high level of correlation of LTC pair co-occurrences in the >65 years and >40 year suggest survival bias has a limited effect (Supplementary Figure 4) 4) Participants may have co-occurring LTCs because of misdiagnosis or codes associated with the diagnostic pathway, for example, the strong co-occurrence observed for polymyalgia rheumatica and fibromyalgia. 5) Completeness of medical records depends on access to healthcare, therefore our analyses may underrepresent communities with less access to services which are less likely to be diagnosed with LTCs. 6) Finally, differences in statistical power for defining conditions as genetically correlated or co-occurring means it is important to consider 95% confidence intervals when considering specific pairs. However, in this study we have used extremely large datasets, ensuring tight confidence intervals around our estimates.

### How could these results help clinicians?

Understanding novel associations and associations involving treatable conditions can highlight opportunities for improved detection, and interventions for prevention, delaying onset or treatment of LTC pairs. Genetic correlations provide a starting point for the identification of specific mechanisms of MLTC providing a foundation for research on potential prevention and treatment.

This knowledge can lead to novel treatment approaches and drug repurposing across LTC pairs that will inform clinical guidelines to the benefit of patients,^32^ or highlight pathways leading to adverse treatment effects. For example, in rheumatoid arthritis and skin cancer our genetic analysis highlighted CTLA4, a shared target for medications across both conditions. This supports growing evidence that immunosuppressants such as abatacept (prescribed to manage rheumatoid arthritis) may increase risk of skin cancers,^33^ and shows the potential of in-depth follow-on analyses of our results.

LTC pairs that are genetically correlated without observed co-occurrence could highlight underdiagnosed conditions, some of which may be amenable to screening or education to improve detection. This includes conditions with a potentially high symptom burden in groups in whom there are barriers to clinical presentation or accessing healthcare, such as people with mental health diagnoses, or highlights conditions that are commonly not diagnosed, such as AAA suggesting screening programs extended only to men over 65 as a one-time event have not been appropriately adopted.^34^

Lastly, we highlight potentially treatable conditions with high co-occurrence, such as iron deficiency anaemia with peripheral neuropathy, and B12 deficiency with diabetes complications and treatments or supplementation advice could be explored further.^35^

### Further work

Future detailed work is planned to investigate the specific relationships between LTC pairs. Multimorbidity represents complex interactions of biological pathways and environmental factors. Longitudinal research, adjusted for confounders, can elucidate mechanisms and risk factors involved for selected, under-researched LTC pairs with strong genetic correlation. Genetic causal inference methodologies can identify targets for intervention,^36^ allowing researchers to test and propose personalised preventative and therapeutic actions.

## Conclusion

We have performed a systematic analysis of multimorbidity, integrating large scale primary-care and genetic data from multiple sources, and involving patients as collaborators. We have identified novel combinations of conditions, including those that tend to share genetic factors but not co-occur in primary-care, and vice versa. Our data is accessible through an interactive web app (https://gemini-multimorbidity.shinyapps.io/atlas), which we anticipate will provide a valuable resource for further research in multimorbidity.

## Data Availability

We cannot make individual-level data available. Researchers can apply to UK Biobank (https://www.ukbiobank.ac.uk/enable-your-research/), CPRD (https://www.cprd.com/research-applications), and SIDIAP (https://www.sidiap.org/index.php/en/solicituds-en). We have made our diagnostic code lists, code and results available on our GitHub (https://github.com/GEMINI-multimorbidity/) site and Shiny website (https://gemini-multimorbidity.shinyapps.io/atlas/). GWAS summary statistics will be available following acceptance at the GWAS Catalog (https://www.ebi.ac.uk/gwas/home) and Zenodo (link on our GitHub [https://github.com/GEMINI-multimorbidity/]).

## Author contributions

All authors read and approved the final version of the manuscript

## Conceptualization

OM, NM, JMV, FD, SEL, JB, DM, JAHM, LCP, JD. Data access and verification: OM, NM, BV, LT, CGM, FD, JB, TMF, LCP, CV, JD. Formal Analysis: OM, NM, BV, LT, CG, AR, JB, JAHM, LCP, JD. Funding Acquisition: CF, LMA, JMV, SK, SEL, MM, LF, KB, JB, DM, TMF, JAHM, LCP, CV, JD. Interpretation: OM, NM, BV, CF, LMA, RMW, XL, JMV, SK, FD, SEL, MM, LF, KB, JB, DM, TMF, JAHM, LCP, CV, JD. Investigation: OM, NM, BV, JB, DM, TMF, JAHM, LCP, CV, JD. Methodology: OM, NM, BV, RMW, XL, JMV, FD, MM, LF, KB, JB, DM, TMF, JAHM, LCP, CV, JD. Patient And Public Involvement: MM, LF, KB. Resources: OM, NM, BV, JAHM, LCP, JD. Software: OM, NM, BV, JB, DM, JAHM, LCP, JD. Supervision: LAC, FD, JB, DM, TMF, JAHM, LCP, CV, JD. Validation: OM, NM, BV, TMF, JAHM, LCP, CV, JD. Visualization: OM, NM, BV, TMF, JAHM, LCP, CV, JD. Writing – Original Draft: OM, NM, BV, TMF, JAHM, LCP, CV, JD. Writing – Review & Editing: OM, BV, LAC, CF, LMA, RMW, XL, JMV, SK, FD, SEL, KB, JB, DM, TMF, JAHM, LCP, CV, JD.

## Acknowledgements

Data from Clinical Practice Research Datalink (CPRD) was obtained under licence from the UK Medicines and Healthcare products Regulatory Agency. The data is provided by patients and collected by the NHS as part of their care and support. The interpretation and conclusions contained in this study are those of the author/s alone. This research has been conducted using the UK Biobank Resource under Application Number 9072. We want to acknowledge the participants and investigators of the FinnGen study. We also thank the authors and study participants for the published GWAS consortium meta-analyses utilized in this report: see the Supplementary Information for full citations. The authors would like to acknowledge the use of the University of Exeter High-Performance Computing (HPC) facility in carrying out this work. For the purpose of open access, the author has applied a Creative Commons Attribution (CC BY) licence to any Author Accepted Manuscript version arising.

## References

1 Kingston A, Robinson L, Booth H, Knapp M, Jagger C. Projections of multi-morbidity in the older population in England to 2035: estimates from the Population Ageing and Care Simulation (PACSim) model. Age Ageing 2018; 47: 374–80.

2 Barnett K, Mercer SW, Norbury M, Watt G, Wyke S, Guthrie B. Epidemiology of multimorbidity and implications for health care, research, and medical education: a cross-sectional study. The Lancet 2012; 380: 37–43.

3 Carrasco-Ribelles LA, Roso-Llorach A, Cabrera-Bean M, et al. Dynamics of multimorbidity and frailty, and their contribution to mortality, nursing home and home care need: A primary care cohort of 1 456 052 ageing people. EClinicalMedicine 2022; 52: 101610.

4 Ho IS-S, Azcoaga-Lorenzo A, Akbari A, et al. Examining variation in the measurement of multimorbidity in research: a systematic review of 566 studies. Lancet Public Health 2021; 6: e587–97.

5 Nichols L, Taverner T, Crowe F, et al. In simulated data and health records, latent class analysis was the optimum multimorbidity clustering algorithm. J Clin Epidemiol 2022; 152: 164–75.

6 Zhu Y, Edwards D, Mant J, Payne RA, Kiddle S. Characteristics, service use and mortality of clusters of multimorbid patients in England: a population-based study. BMC Med 2020; 18: 78.

7 Dong G, Feng J, Sun F, Chen J, Zhao X-M. A global overview of genetically interpretable multimorbidities among common diseases in the UK Biobank. Genome Med 2021; 13: 110.

8 Kim S-S, Hudgins AD, Gonzalez B, et al. A Compendium of Age-Related PheWAS and GWAS Traits for Human Genetic Association Studies, Their Networks and Genetic Correlations. Front Genet 2021; 12. DOI:10.3389/fgene.2021.680560.

9 West CE, Karim M, Falaguera MJ, et al. Integrative GWAS and co-localisation analysis suggests novel genes associated with age-related multimorbidity. Sci Data 2023; 10: 655.

10 Bulik-Sullivan BK, Loh P-R, Finucane HK, et al. LD Score regression distinguishes confounding from polygenicity in genome-wide association studies. Nat Genet 2015; 47: 291–5.

11 Bulik-Sullivan B, Finucane HK, Anttila V, et al. An atlas of genetic correlations across human diseases and traits. Nat Genet 2015; 47: 1236–41.

12 Wolf A, Dedman D, Campbell J, et al. Data resource profile: Clinical Practice Research Datalink (CPRD) Aurum. Int J Epidemiol 2019; 48: 1740–1740g.

13 Recalde M, Rodríguez C, Burn E, et al. Data Resource Profile: The Information System for Research in Primary Care (SIDIAP). Int J Epidemiol 2022; 51: e324–36.

14 Sudlow C, Gallacher J, Allen N, et al. UK Biobank: An Open Access Resource for Identifying the Causes of a Wide Range of Complex Diseases of Middle and Old Age. PLoS Med 2015; 12: e1001779.

15 Kurki MI, Karjalainen J, Palta P, et al. FinnGen provides genetic insights from a well-phenotyped isolated population. Nature 2023; 613: 508–18.

16 Sollis E, Mosaku A, Abid A, et al. The NHGRI-EBI GWAS Catalog: knowledgebase and deposition resource. Nucleic Acids Res 2023; 51: D977–85.

17 Calderón-Larrañaga A, Vetrano DL, Onder G, et al. Assessing and Measuring Chronic Multimorbidity in the Older Population: A Proposal for Its Operationalization. J Gerontol A Biol Sci Med Sci 2017; 72: 1417–23.

18 Kuan V, Denaxas S, Gonzalez-Izquierdo A, et al. A chronological map of 308 physical and mental health conditions from 4 million individuals in the English National Health Service. Lancet Digit Health 2019; 1: e63–77.

19 Mbatchou J, Barnard L, Backman J, et al. Computationally efficient whole-genome regression for quantitative and binary traits. Nat Genet 2021; 53: 1097–103.

20 Dettori JR, Norvell DC, Chapman JR. Fixed-Effect vs Random-Effects Models for Meta-Analysis: 3 Points to Consider. Global Spine J 2022; 12: 1624–6.

21 Mägi R, Morris AP. GWAMA: software for genome-wide association meta-analysis. BMC Bioinformatics 2010; 11: 288.

22 Giambartolomei C, Vukcevic D, Schadt EE, et al. Bayesian Test for Colocalisation between Pairs of Genetic Association Studies Using Summary Statistics. PLoS Genet 2014; 10: e1004383.

23 Piñero J, Ramírez-Anguita JM, Saüch-Pitarch J, et al. The DisGeNET knowledge platform for disease genomics: 2019 update. Nucleic Acids Res 2019; published online Nov 4. DOI:10.1093/nar/gkz1021.

24 Knox C, Wilson M, Klinger CM, et al. DrugBank 6.0: the DrugBank Knowledgebase for 2024. Nucleic Acids Res 2024; 52: D1265–75.

25 GEMINI. GEMINI - Get Involved. 2021. https://sites.exeter.ac.uk/gemini/get-involved/ (accessed Jan 20, 2024).

26 Mounier N, Voller B, Masoli JA, et al. Genetics identifies obesity as a shared risk factor for co-occurring multiple long-term conditions. Corresponding author. medRxiv 2024. DOI:10.1101/2024.07.10.24309772.

27 Sodini SM, Kemper KE, Wray NR, Trzaskowski M. Comparison of Genotypic and Phenotypic Correlations: Cheverud’s Conjecture in Humans. Genetics 2018; 209: 941–8.

28 Defeudis G, Mazzilli R, Tenuta M, et al. Erectile dysfunction and diabetes: A melting pot of circumstances and treatments. Diabetes Metab Res Rev 2022; 38. DOI:10.1002/dmrr.3494.

29 Nirwan JS, Hasan SS, Babar Z-U-D, Conway BR, Ghori MU. Global Prevalence and Risk Factors of Gastro-oesophageal Reflux Disease (GORD): Systematic Review with Meta-analysis. Sci Rep 2020; 10: 5814.

30 Momen NC, Plana-Ripoll O, Agerbo E, et al. Association between Mental Disorders and Subsequent Medical Conditions. New England Journal of Medicine 2020; 382: 1721–31.

31 Kugathasan P, Stubbs B, Aagaard J, Jensen SE, Munk Laursen T, Nielsen RE. Increased mortality from somatic multimorbidity in patients with schizophrenia: a Danish nationwide cohort study. Acta Psychiatr Scand 2019; 140: 340–8.

32 Tan GSQ, Sloan EK, Lambert P, Kirkpatrick CMJ, Ilomäki J. Drug repurposing using real-world data. Drug Discov Today 2023; 28: 103422.

33 Kreher MA, Konda S, Noland MMB, Longo MI, Valdes-Rodriguez R. Risk of melanoma and nonmelanoma skin cancer with immunosuppressants, part II: Methotrexate, alkylating agents, biologics, and small molecule inhibitors. J Am Acad Dermatol 2023; 88: 534–42.

34 Benson RA, Meecham L, Fisher O, Loftus IM. Ultrasound screening for abdominal aortic aneurysm: current practice, challenges and controversies. Br J Radiol 2018; 91: 20170306.

35 Aroda VR, Edelstein SL, Goldberg RB, et al. Long-term Metformin Use and Vitamin B12 Deficiency in the Diabetes Prevention Program Outcomes Study. J Clin Endocrinol Metab 2016; 101: 1754– 61.

36 Masoli JAH, Pilling LC, Frayling TM. Genomics and multimorbidity. Age Ageing 2022; 51. DOI:10.1093/ageing/afac285.

