## Supplementary Information for "A systematic analysis of the contribution of genetics to multimorbidity and comparisons with primary care data"

**Strobe Checklist**

|  | **Item No** | **Recommendation** | **Page No** |
| --- | --- | --- | --- |
| **Title and abstract** | 1 | (*a*) Indicate the study’s design with a commonly used term in the title or the abstract | 1 |
|  |  | (*b*) Provide in the abstract an informative and balanced summary of what was done and what was found | 2 |
| **Introduction** | | | |
| Background/rationale | 2 | Explain the scientific background and rationale for the investigation being reported | 3,5, |
| Objectives | 3 | State specific objectives, including any prespecified hypotheses | 5 |
| **Methods** | | | |
| Study design | 4 | Present key elements of study design early in the paper | 5,6,7 |
| Setting | 5 | Describe the setting, locations, and relevant dates, including periods of recruitment, exposure, follow-up, and data collection | 5, Supplementary Methods |
| Participants | 6 | (*a*) Give the eligibility criteria, and the sources and methods of selection of participants | 5, Supplementary Methods |
| Variables | 7 | Clearly define all outcomes, exposures, predictors, potential confounders, and effect modifiers. Give diagnostic criteria, if applicable | Supplementary Methods |
| Data sources/ measurement | 8* | For each variable of interest, give sources of data and details of methods of assessment (measurement). Describe comparability of assessment methods if there is more than one group | *Supplementary Methods* |
| Bias | 9 | Describe any efforts to address potential sources of bias | Corrected for age and gender, multiple testing 7 |
| Study size | 10 | Explain how the study size was arrived at | Supplementary Methods,6 |
| Quantitative variables | 11 | Explain how quantitative variables were handled in the analyses. If applicable, describe which groupings were chosen and why | Age: Supplementary methods |
| Statistical methods | 12 | (*a*) Describe all statistical methods, including those used to control for confounding | 7 |
|  |  | (*b*) Describe any methods used to examine subgroups and interactions | NA |
|  |  | (*c*) Explain how missing data were addressed | NA |
|  |  | (*d*) If applicable, describe analytical methods taking account of sampling strategy | NA |
|  |  | (*e*) Describe any sensitivity analyses | NA |
| **Results** | | | |
| Participants | 13* | (a) Report numbers of individuals at each stage of study—eg numbers potentially eligible, examined for eligibility, confirmed eligible, included in the study, completing follow-up, and analysed | Supplementary methods 1 |
|  |  | (b) Give reasons for non-participation at each stage | Supplementary Methods |
|  |  | (c) Consider use of a flow diagram | Used for diseases not participants (Supplementary Methods) |
| Descriptive data | 14* | (a) Give characteristics of study participants (eg demographic, clinical, social) and information on exposures and potential confounders | Supplementary table 2 |
|  |  | (b) Indicate number of participants with missing data for each variable of interest | NA |
| Outcome data | 15* | Report numbers of outcome events or summary measures | Shiny app |
| Main results | 16 | (*a*) Give unadjusted estimates and, if applicable, confounder-adjusted estimates and their precision (eg, 95% confidence interval). Make clear which confounders were adjusted for and why they were included | Supplementary table 4-5 |
|  |  | (*b*) Report category boundaries when continuous variables were categorized | Table 1 page 20 |
|  |  | (*c*) If relevant, consider translating estimates of relative risk into absolute risk for a meaningful time period | NA |
| Other analyses | 17 | Report other analyses done—eg analyses of subgroups and interactions, and sensitivity analyses | NA |
| **Discussion** | | | |
| Key results | 18 | Summarise key results with reference to study objectives | 11 |
| Limitations | 19 | Discuss limitations of the study, taking into account sources of potential bias or imprecision. Discuss both direction and magnitude of any potential bias | 12 |
| Interpretation | 20 | Give a cautious overall interpretation of results considering objectives, limitations, multiplicity of analyses, results from similar studies, and other relevant evidence | 11-12 |
| Generalisability | 21 | Discuss the generalisability (external validity) of the study results | 12 |
| **Other information** | | | |
| Funding | 22 | Give the source of funding and the role of the funders for the present study and, if applicable, for the original study on which the present article is based | 14 |

### Research in Context

#### Literature Review

We searched PUBMED, Embase, Global Health for articles published in English, since 6. Date >2012 (after the definitive Barnett et al. multimorbidity review paper was published) until 01 May 2024, using the search terms: (((multidisease*[tiab] OR multi-disease*[tiab] OR multimorbidit*[tiab] OR multi-morbidit*[tiab] OR multipatholog*[tiab] OR multi-patholog*[tiab] OR pluripatholog*[tiab] OR polypatholog*[tiab] OR poly-pathology*[tiab] OR "multiple long term conditions"[tiab])) OR (multimorbidity[MeSH Terms])) AND ("genetic*"[tiab] OR "genomic*"[tiab] OR ("genome wide association stud*"[tiab] OR "genome wide association analysis"[tiab] OR "GWAS"[tiab])) NOT (animals[mh] NOT humans[mh]) AND NOT ("retracted publication"[pt]). We excluded studies investigating monogenic mutations and case studies. 243 studies met the primary inclusion criteria, where 68 presented original research addressing multimorbidity and incorporating genetic analysis. Multimorbidity is predominantly characterised by counts or clusters of conditions, with limited exploration of underlying mechanisms. Only 11 investigated multimorbidity beyond the analysis of comorbidities of single conditions, with five reporting systematic investigations of genetics across multiple long-term conditions.^1–5^

### Supplementary Methods

#### Disease Code lists

We have adopted definitions used in previous projects and published in peer-reviewed journals with additional curation by clinical fellows. This provides confidence that such criteria for defining exposures and outcomes are sound, minimising the potential effect of misclassification in produced estimates.

Code lists for the 84 diseases are available from:

<https://github.com/GEMINI-multimorbidity/GEMINI-LTC-code-list-Public>

Codelist by condition: <https://github.com/GEMINI-multimorbidity/GEMINI-LTC-code-list-Public/tree/main/codelist%20by%20condition>

#### Data sources

##### Electronic healthcare records: CPRD and SIDIAP

Two datasets, the UK Clinical Practice Research Data Link (CPRD) AURUM and The Information System for Research in Primary Care (SIDIAP) in Spain, were used for a cross-sectional analysis of diagnoses in these routine health records are coded to standardise clinical diagnoses and contribute to disease registers and healthcare planning.

CPRD AURUM data captures demographic, diagnostic, drug, lab and referral information from patients attending 1,491 UK-based GPs, distributed across England. The data is regularly collected and updated with de-identified data made available to researchers upon acceptance of a study protocol. The extract contains several files including Observation, Drug Issue, Patient, Staff, Practice, Referral and Consultation. This dataset includes data on over 40 million patients, of which 13,300,067, 19.8% of the English population are currently alive and enrolled with a practice. Patients included in CPRD AURUM are broadly representative of the English population. Diagnosis in CPRD are recorded using Read Codes v2, EMIS codes and SNOMED CT codes. In this study, we included N=2,425,014 patients alive, registered and over the age of 65 on 01/01/2020. Patients were considered to have a disease with at least one observation matching the code list before 01/01/2020.

SIDIAP captures demographic, diagnostic, drug, lab and referral information from patients attending 328 primary-care centres that cover 75% of the population living in Catalonia, Spain. The database is updated every 6 months and is structured in data domains, each containing the person’s pseudo-anonymized identifier. Data is accessible to all researchers following approval of a research protocol. The extract contains information on Socio-demographics, Health conditions, Medications and vaccines, Laboratory tests, Clinical practice and lifestyle information, and Sexual and reproductive health. The database has information on 8,036,948 people, of whom 5,801,280 (72.2%) were still active as of 30 June 2021. SIDIAP is representative of the population of Catalonia in terms of age, sex and geographic distribution. SIDIAP the electronic codes are recorded using the International Classification of Disease, version 10 (ICD-10-CM/PCS).^6^ In this study, we included N= 1,053,640 patients alive, registered and over the age of 65 on 01/01/2020 were analysed. Patients were considered to have a disease with at least one observation matching the code list before 01/01/2020.

##### Genetic Data

Genetic data were collected from three different sources: 1) UK Biobank [UKB],^7^ a large population-based prospective study with 450,197 individuals of European genetic ancestry. 2) FinnGen, a large-scale genomics initiative including over 500,000 participants with linked health diagnosis data. 3) Disease-specific GWAS meta-analyses summary statistics when available for each LTC.

##### UK Biobank data

To perform the genetic analyses we ascertained diagnosis of LTCs using both primary-care linked data (available for 45% of participants, censoring date: 28/02/2016 – Read v2 and CTV3 codes, truncated to 5 bytes) and hospital inpatient diagnoses (available for all participants, censoring date: 31/10/2022 - ICD-10 codes). Participants were genotyped using two near identical (>95% shared variants, n=805,426 total) microarray platforms: the Affymetrix Axiom UK Biobank array (in 438,427 participants) and the Affymetrix UKBiLEVE array (in 49,950 participants). UK Biobank centrally performed genotype imputation in 487,442 participants using data from the Haplotype Reference Consortium and UK10K reference panels, increasing the number of genetic variants to ~96 million.^8^ We exclude genetic variants with <0.1% minor allele frequency or with imputed INFO score <0.3, leaving ~16 million for GWAS analysis. GWAS were performed in up to 451,197 participants genetically similar to the 1000 Genomes EUR population (described previously.^9^ In brief, individuals from the UK Biobank were projected into the 1000 Genomes principal component (PC) space using the SNP loadings derived from the initial PC analysis to minimise confounding of PC values due to varying degrees of relatedness within UK Biobank.^10^ Using the means derived from the 1000 Genomes reference dataset, we subsequently performed K-means clustering analyses to determine which individuals from UK Biobank could be classified as EUR-like. GWAS were performed in UKB participants genetically similar to the 1000 Genomes EUR reference population for 84 LTCs, using the same clinical code lists as above in CPRD, using the REGENIE software (v3.1.3) to account for population structure and relatedness, adjusted for age at baseline assessment, sex, genotyping chip, and assessment centre. ^11^ For quality control, we restricted variants to those with a minor allele frequency (MAF) of >0.1%, and an imputation INFO score ≥0.3.

##### FinnGen data

FinnGen is a large-scale genomics initiative, that contains data from over 500,000 participants and is linked to health diagnosis data. GWAS summary statistics from the FinnGen cohort (release 9) with 377,277 participants, provided for predetermined disease (“endpoints”), defined using ICD-10-FM (Finnish Modification). ^12^

##### Disease-specific GWAS

Disease-specific GWAS meta-analyses summary statistics when available for each LTC. We used the GWAS Catalog (<https://www.ebi.ac.uk/gwas>), ^13^ disease-specific public repositories and contacted authors of the latest GWAS to identify relevant studies with aligned disease definitions and participants of European ancestry to enable comparison with UKB and FinnGen. The below LTCs had available published and available GWAS summary statistics and were used in the genetics analysis (see Supplementary Table 1 for further information).

- Anxiety disorders.^14^
- Asthma.^15^
- Atrial fibrillation.^16^
- Chronic kidney disease.^17^
- Chronic obstructive pulmonary disease.^18^
- Coronary heart disease.^19^
- Depression.^20^
- Erectile dysfunction.^21^
- Gastro-oesophageal reflux disease.^22^
- Glaucoma.^23^
- Gout.^24^
- Hearing loss.^25^
- Heart failure.^26^
- Hyperthyroidism, hypothyroidism.^27^
- Irritable bowel syndrome.^28^
- Migraine.^29^
- Osteoarthritis.^30^
- Primary breast malignancy.^31^
- Rheumatoid arthritis.^32^
- Schizophrenia, schizotypal and delusional disorders.^33^
- Type 2 diabetes.^34^
- Ulcerative colitis.^35^

##### Defining Long-Term Conditions

In GEMINI, Long-Term Conditions (LTCs) are defined by 3 criteria: 1) Chronicity, conditions with a course typically lasting longer than 3 months or producing sequelae lasting more than 3 months. 2) Prevalence, LTCs with a prevalence greater than 0.5% in a population aged 65 and over. 3) Genetic heritability: we selected LTCs with a statistically significant common genetic component (see below).

##### Step 1 (Selection of conditions – chronicity)

A set of 308 conditions and respective clinical coding lists (ICD-10 and Read v2) are available from the CALIBER platform, a publicly available resource with code lists for medical conditions.^36^ Disease chronicity was defined by the “Karolinska” list (KL) for the classification of chronic morbidity. ^37^ KL details ICD-10 codes that represent chronic conditions, with codes excluded from KL interpreted as acute and out of scope. We compared the CALIBER code lists against the KL. N=144 CALIBER lists matching perfectly with KL were classified as LTCs and taken forward. N=88 CALIBER lists match partially with KL, presenting a mixture of chronic and acute codes. For partial matching code lists, two clinicians (JM and CV) were tasked with revising code lists independently and identifying Read v2 and ICD10 codes that refer to acute conditions. Acute codes were then marked for exclusion. A third clinician (DM) resolved conflicts. N=62 CALIBER lists had no matching codes with KL and were excluded. N=14 CALIBER lists were excluded as all codes were already included in other CALIBER lists. Figure 1 is a flowchart of condition selection for chronicity, prevalence, and heritability.

##### Step 2: Selection of conditions – prevalence

Prevalence for all conditions was estimated using CPRD data for any individuals aged 65 and older who were alive and registered with a practice on the 1^st^ of January 2020. The cutoff date minimises the impact of the COVID-19 pandemic in our analyses. LTCs with a prevalence under 0.5% in participants ages 65 years and older in either CPRD or SIDIAP were excluded to ensure adequate statistical power when studying specific LTC combinations in downstream analyses. A Patient and Public Involvement (PPI) workshop reviewed the initial code lists for relevance and importance with N=28 additions. A total of N=84 (40 where CALIBER and KL were a perfect match and 44 revised by clinicians) LTC code lists were selected for heritability analysis.

##### Step 3: Selection of conditions - Heritability

Heritability measures the proportion of phenotypic variance explained by genetics. Based on UKB GWAS summary statistics, we estimated the SNP-based heritability – the proportion of phenotypic variance explained by additive effects of a set of SNPs – using LD-score regression (LDSC).^38,39^ We used the provided EUR reference population LD data throughout. LTCs with a heritability estimate Z-score >4 were taken forward in the analysis.^40^

**Co-occurrence of conditions in primary care electronic records**

Logistic regression models identified co-occurring LTCs (MLTC) in observational data: adjusted for age (continuous measurement) and gender and a Benjamini-Hochberg correction was used to account for multiple tests. The correlation between LTCs estimated in CPRD AURUM and SIDIAP datasets was meta-analysed, with fixed effects using the RMA function in the R package ‘metafor’. Fixed effects models are appropriate when few data sources are available (N=2) (insert PMID: 35723546).

The Logistic regression results were adjusted for age and gender. The logistic regression outcomes were adjusted for age and gender. Consequently, the odds ratios exhibit asymmetry when the constituent LTCs of a pair are interchanged as either "outcome" or "exposure." For the representation of the results from observational data, we selected the direction of the analysis that showed the least heterogeneity between SIDIAP and CPRD based on the q-statistic of the meta-analysis.

**GWAS Meta-analysis and Genetic Correlation**

For the 72 conditions meeting the heritability criteria above, we meta-analysed genome-wide summary data from up to 3 data sources – UKB, FinnGen and disease-specific GWAS (referred to as Consortium data). See the Supplementary Methods for individual study references, Supplementary Figure 2 for analysis flowchart, and Supplementary Table 1 for effective sample size and other information. A cross-trait LD-score regression framework, that estimates the within-condition, between-dataset genetic correlation, measured the similarity between conditions. ^40^ The FinnGen and Consortium data were added to the meta-analysis when within-condition genetic correlation ($R_{g}$) with UK Biobank was >0.8. Where consortium data included UK Biobank or FinnGen data, the consortium data was used to avoid overlapping datasets (i.e., if UKB was in the consortium GWAS, then we only meta-analysed consortium+FinnGen). Studies were meta-analysed using GWAMA. ^41^

The meta-analysed GWAS summary statistics and LD-score regression identified pairwise genetic correlations between the 72 LTCs,^40^ with significance set at a false discovery rate (FDR) of 5%.

2556 pairs were reduced to 2546 pairs for clinical review and for the scatter plot as 10 pairs had partially overlapping code lists which gave rise to extremely high odds ratios (e.g stroke and TIA).

LTC pairs were distinguished within and across domain and separated into terciles based on within-domain MLTCs: (1) low (2) medium and (3) high genetic correlation.

### Supplementary Figures

#### SFigure 1: Flowchart of conditions selection for chronicity, prevalence, and heritability

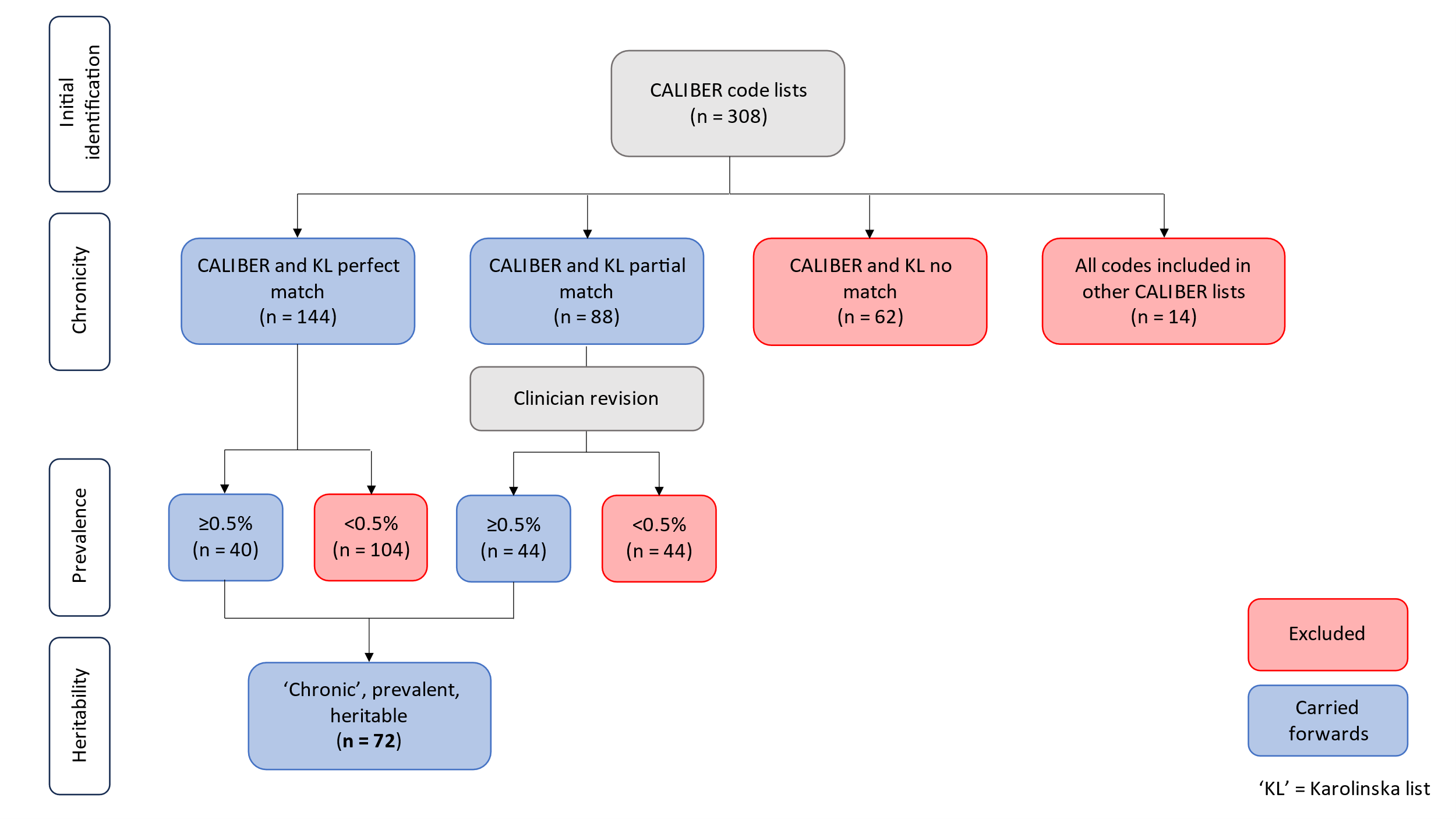

#### SFigure 2: Flowchart for data sources for GWAS meta-analysis

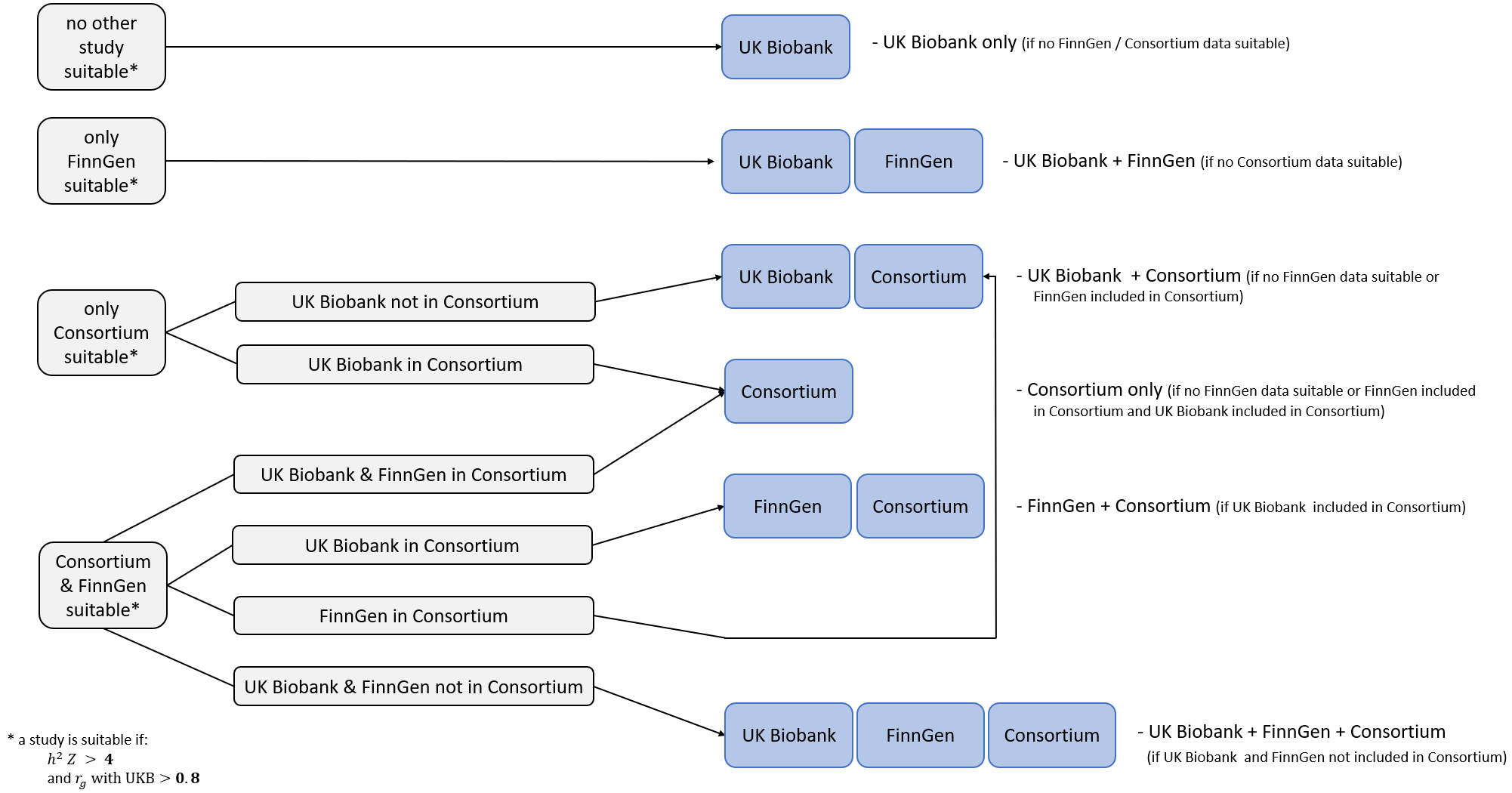

#### SFigure 3: The relationship between LTC observational co-occurrence and genetic correlation (stratified by domain)

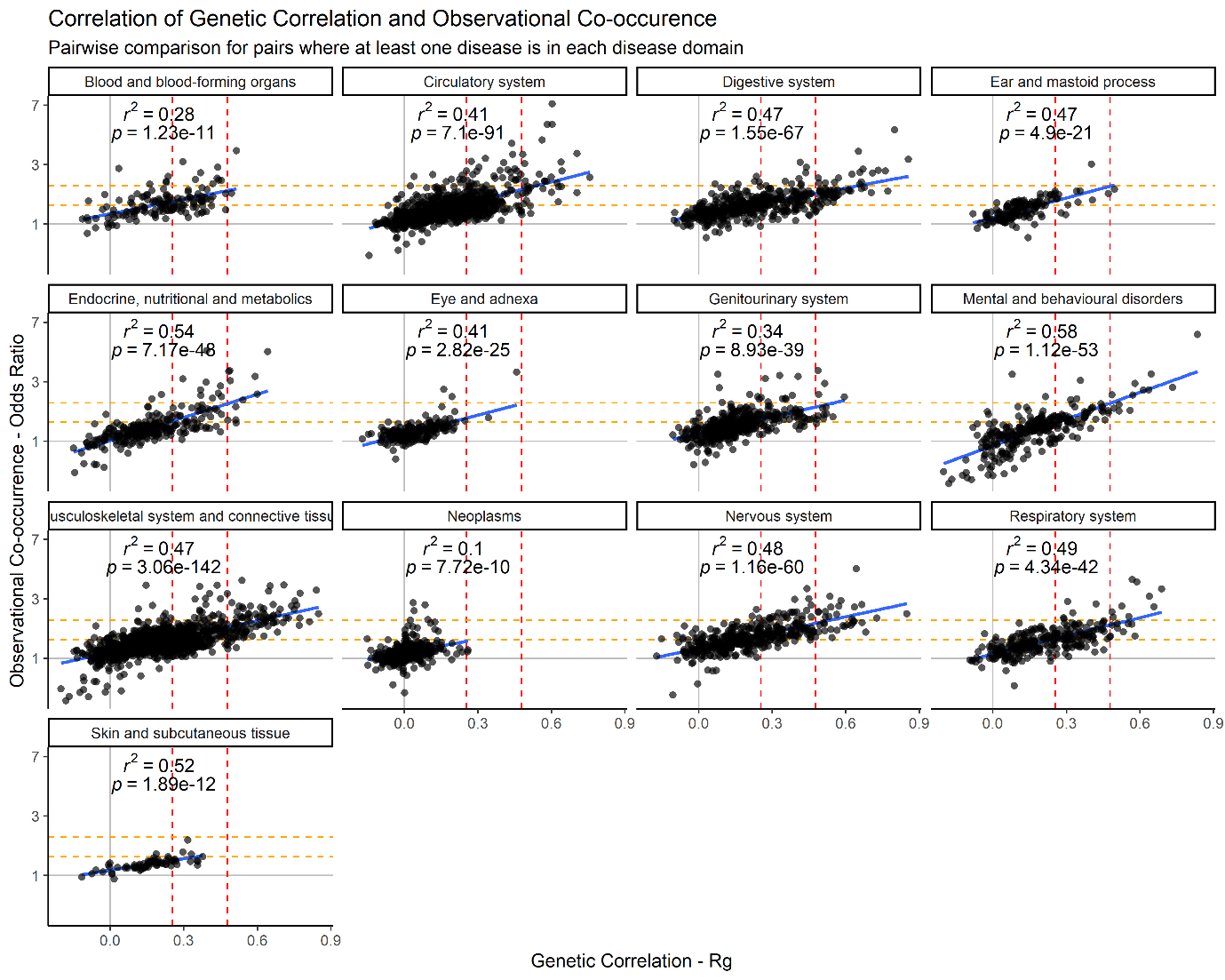

*Scatter plots of the relationship between observed co-occurrence and genetic correlation of LTC pairs. (blue) linear regression line, (yellow) terciles of genetic correlation, (red) terciles of likelihood of observed cooccurrence. Terciles estimated based on within-domain LTC pairs.*

**SFigure 4: Sensitivity analysis of CPRD only log odds ratios in patients 65 years and over (the main focus of the paper) versus log odds ratios in patients over 40 years of age.**

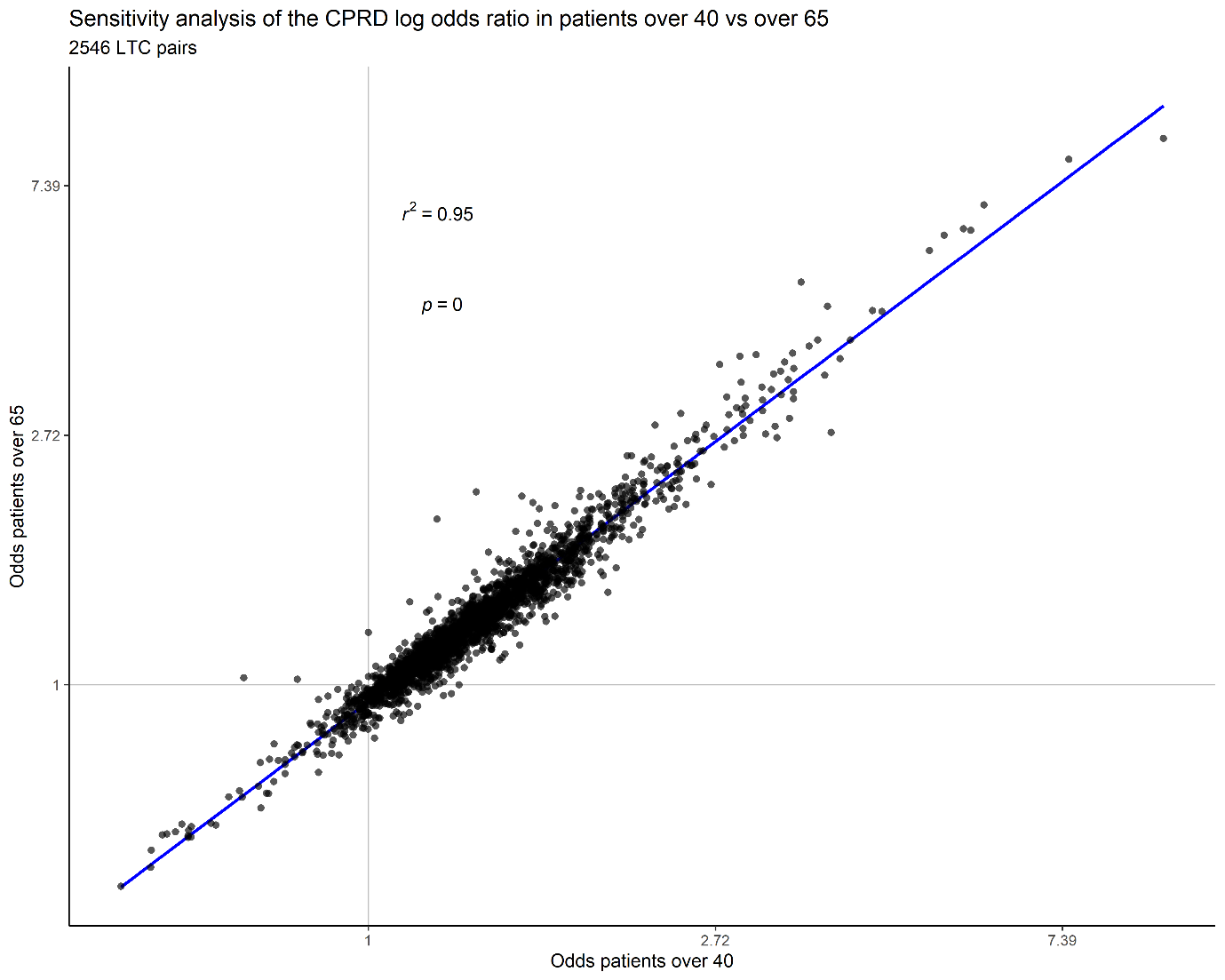

*CPRD only log odds of 2546 LTC pairs adjusted for age and gender for patients over 40 versus patients over 60, the results were highly correlated.*

### Supplementary Tables

Supplementary tables are available on the GitHub repositories (<https://github.com/GEMINI-multimorbidity/Paper1-Supplementary-Tables>)

Interactive versions are also available in our shiny app: <https://gemini-multimorbidity.shinyapps.io/atlas>.

**STable 1: A summary of information of 84 Gemini diseases, icd domain classification, prevalence in observational datasets, heritability , GWAS effective sample size and data sources**

<https://raw.githubusercontent.com/GEMINI-multimorbidity/Paper1-Supplementary-Tables/refs/heads/main/githubGEMINI%20Atlas%20paper%20-%20Supplementary%20Tables_ST1.csv>
**STable 2: A table of the population characteristics in the datasets accessed (CPRD, SIDIAP and UK BIOBANK) Age and diagnosis at index date (2020 for health record data and 2016 (last available data collection date) in UK Biobank. The average number of GEMINI conditions out of 72 at index date is calculated.**

| Category | SIDIAP (2020) | CPRD (2020) | UK Biobank (2016) |
| --- | --- | --- | --- |
| N | 1,053,640 | 2,425,014 | 451,240 |
| Age at Index |  |  |  |
| 65-74 | 523,708 (49·7) | 1,264,804 (52·2) | 220,689 (83·0) |
| 75-84 | 347,858 (33) | 798,554 (32·9) | 45,082 (17·0) |
| 85+ | 182,074 (17·3) | 361,656 (14·9) | 0 (0) |
| gender |  |  |  |
| Male | 452,608 (43) | 1,121,914 (46·3) | 206,377 (45·7) |
| Female | 601,032 (57) | 1,303,100 (53·7) | 244,863 (54·3) |
| N GEMINI conditions (out of 72) | |  |  |
| 0 | 29,309 (2·8) | 101,343 (4·2) | 78,951 (17·5) |
| 1 | 53,104 (5) | 146,650 (6·0) | 56,521 (12·5) |
| 2 | 81,514 (7·7) | 194,493 (8·0) | 53,054 (11·8) |
| 3 | 10,5340 (10) | 228,549 (9·4) | 48,611 (10·8) |
| 4 | 12,1459 (11·5) | 242,893 (10·0) | 41,798 (9·3) |
| 5 | 12,5747 (11·9) | 242,235 (10·0) | 35,479 (7·9) |
| 6 | 12,0043 (11·4) | 229,808 (9·5) | 29,313 (6·5) |
| 7 | 10,6829 (10·1) | 206,303 (8·5) | 23,897 (5·3) |
| 8 | 89,373 (8·5) | 177,795 (7·3) | 19,223 (4·3) |
| 9 | 69,606 (6·6) | 150,519 (6·2) | 14,949 (3·3) |
| 10 | 51,954 (4·9) | 123,063 (5·1) | 12,100 (2·7) |
| 11 | 36,266 (3·4) | 97,819 (4·0) | 9,397 (2·1) |
| 12 | 24,527 (2·3) | 75,940 (3·1) | 7128 (1·6) |
| 13-15 | 31,302 (3) | 133,759 (5·5) | 12,726 (2·8) |
| 16-20 | 7043 (0·7) | 64,228 (2·6) | 6,713 (1·5) |
| 21+ | 224 (0) | 9617 (0·4) | 1380 (0·3) |

**STable 3: Number of diseases assigned to 13 ICD-10 chapter domains for the final 72 conditions**

| ICD-10 domain | Number of LTCs |
| --- | --- |
| Diseases of the circulatory system | 12 |
| Diseases of the genitourinary system | 6 |
| Diseases of the respiratory system | 4 |
| Diseases of the digestive system | 7 |
| Diseases of the blood and blood-forming organs and certain disorders involving the immune mechanism | 2 |
| Diseases of the musculoskeletal system and connective tissue | 16 |
| Mental and behavioural disorders | 4 |
| Diseases of the nervous system | 6 |
| Diseases of the eye and adnexa | 3 |
| Diseases of the ear and mastoid process | 2 |
| Endocrine, nutritional and metabolic diseases | 4 |
| Neoplasms | 5 |
| Diseases of the skin and subcutaneous tissue | 1 |
| Total | 72 |

**STable 4: Meta analysed odds ratios for 2556 pairs of diseases.** **The p values are adjusted for multiple testing based on 2556 pairs (p_bh_heatmap) and 2546 pairs p_bh_scatter plot) due to pairs with similar codelists being removed. The Q test was conducted to assess the heterogeneity among effect sizes in the fixed-effects meta-analysis** **The null hypothesis of the Q test is that there is no true heterogeneity among the effect sizes, meaning that any observed differences are due to sampling error alone.**

<https://raw.githubusercontent.com/GEMINI-multimorbidity/Paper1-Supplementary-Tables/refs/heads/main/githubGEMINI%20Atlas%20paper%20-%20Supplementary%20Tables_ST4.csv>

**STable 5: Genetic correlation (Rg) for 2556 diseases pairs.** **The p values are adjusted for multiple testing based on 2556 pairs (p_fdr_heatmap) and 2546 pairs p_fdr_scatter plot) due to pairs with similar codelists being removed.**

<https://raw.githubusercontent.com/GEMINI-multimorbidity/Paper1-Supplementary-Tables/refs/heads/main/githubGEMINI%20Atlas%20paper%20-%20Supplementary%20Tables_ST5.csv>

**STable 6: Results- Mentioned pairs: associations and strength of all pairs mentioned in the results section by section number**

For full table download from: <https://raw.githubusercontent.com/GEMINI-multimorbidity/Paper1-Supplementary-Tables/refs/heads/main/githubGEMINI%20Atlas%20paper%20-%20Supplementary%20Tables_ST6.xlsx>

| ltc 1 | ltc 2 | Results section category | Gen Strength | Obs Strength | OR | Rg | domain |
| --- | --- | --- | --- | --- | --- | --- | --- |
| Atrial fibrillation | Heart failure | 1 | Strong | Strong | OR: 7.53 [7.45-7.61] | Rg=0.60-SE=0.03 | within domain |
| Chronic sinusitis | Gastro-oesophageal reflux disease | 2 | Strong | Strong | OR: 2.00 [1.98-2.02] | Rg=0.49-SE=0.06 | across domain |
| Erectile dysfunction | Peripheral neuropathies (excluding cranial nerve and carpal tunnel syndromes) | 2 | Strong | Strong | OR: 2.20 [2.16-2.23] | Rg=0.49-SE=0.10 | across domain |
| Iron deficiency anaemia | Peripheral arterial disease | 2 | Intermediate | Strong | OR: 2.08 [2.04-2.12] | Rg=0.45-SE=0.10 | across domain |
| Coronary Heart Disease | All stroke | 2 | Strong | Strong | OR: 2.06 [2.04-2.09] | Rg=0.49-SE=0.03 | within domain |
| Rheumatoid Arthritis | Polymyalgia Rheumatica | 2 | Strong | Strong | OR: 2.96 [2.87-3.05] | Rg=0.48-SE=0.09 | within domain |
| Transient ischaemic attack | Peripheral arterial disease | 2 | Intermediate | Strong | OR: 2.30 [2.24-2.35] | Rg=0.48-SE=0.08 | within domain |
| Diabetes type 2 | Erectile dysfunction | 2 | Strong | Strong | OR: 3.27 [3.24-3.30] | Rg=0.49-SE=0.05 | across domain |
| Osteoarthritis (excl spine) | Obesity | 2 | Strong | Strong | OR: 2.00 [1.99-2.01] | Rg=0.54-SE=0.02 | across domain |
| Intervertebral disc disorders | Gastro-oesophageal reflux disease | 2 | Strong | Intermediate | OR: 1.60 [1.58-1.62] | Rg=0.50-SE=0.03 | across domain |
| Tendon disorders | Diverticular disease of intestine (acute and chronic) | 2 | Intermediate | Intermediate | OR: 1.49 [1.48-1.50] | Rg=0.27-SE=0.03 | across domain |
| Fibromyalgia | Irritable bowel syndrome | 2 | Strong | Strong | OR: 3.38 [3.30-3.47] | Rg=0.65-SE=0.13 | across domain |
| Fibromyalgia | Asthma | 2 | Intermediate | Intermediate | OR: 1.87 [1.83-1.92] | Rg=0.45-SE=0.06 | across domain |
| Irritable bowel syndrome | Peripheral neuropathies (excluding cranial nerve and carpal tunnel syndromes) | 2 | Strong | Intermediate | OR: 1.60 [1.57-1.64] | Rg=0.57-SE=0.12 | across domain |
| Vitamin B12 deficiency anaemia | COPD | 2 | Intermediate | Intermediate | OR: 1.57 [1.55-1.60] | Rg=0.35-SE=0.06 | across domain |
| Primary Malignancy Other Skin and subcutaneous tissue | Rheumatoid Arthritis | 3 | Weak | Weak | OR: 1.15 [1.12-1.18] | Rg=-0.14-SE=0.04 | across domain |
| Enthesopathies of the upper body | Schizophrenia, schizotypal and delusional disorders | 3 | Weak | Weak | OR: 0.53 [0.51-0.55] | Rg=-0.17-SE=0.03 | across domain |
| Female genital prolapse | Diabetes type 2 | 4 | Weak | Weak | OR: 0.97 [0.96-0.98] | Rg=0.13-SE=0.02 | across domain |
| Fibromyalgia | Schizophrenia, schizotypal and delusional disorders | 4 | Weak | Weak | OR: 0.84 [0.75-0.94] | Rg=0.20-SE=0.04 | across domain |
| Fibromyalgia | Polymyalgia Rheumatica | 4 | Not Significant | Strong | OR: 3.41 [3.28-3.54] | Rg=0.15-SE=0.12 | within domain |
| Primary Malignancy Colorectal | Iron deficiency anaemia | 4 | Not Significant | Strong | OR: 2.55 [2.49-2.60] | Rg=0.04-SE=0.07 | across domain |
| Primary Malignancy Bladder | Abdominal aortic aneurysm | 4 | Not Significant | Strong | OR: 2.23 [2.09-2.37] | Rg=0.04-SE=0.12 | across domain |

### References

1	Amell A, Roso-Llorach A, Palomero L, *et al.* Disease networks identify specific conditions and pleiotropy influencing multimorbidity in the general population. *Sci Rep* 2018; **8**: 15970.

2	Fadason T, Schierding W, Lumley T, O’Sullivan JM. Chromatin interactions and expression quantitative trait loci reveal genetic drivers of multimorbidities. *Nat Commun* 2018; **9**: 5198.

3	Dong G, Feng J, Sun F, Chen J, Zhao X-M. A global overview of genetically interpretable multimorbidities among common diseases in the UK Biobank. *Genome Med* 2021; **13**: 110.

4	Kim S-S, Hudgins AD, Gonzalez B, *et al.* A Compendium of Age-Related PheWAS and GWAS Traits for Human Genetic Association Studies, Their Networks and Genetic Correlations. *Front Genet* 2021; **12**. DOI:10.3389/fgene.2021.680560.

5	West CE, Karim M, Falaguera MJ, *et al.* Integrative GWAS and co-localisation analysis suggests novel genes associated with age-related multimorbidity. *Sci Data* 2023; **10**: 655.

6	Recalde M, Rodríguez C, Burn E, *et al.* Data Resource Profile: The Information System for Research in Primary Care (SIDIAP). *Int J Epidemiol* 2022; **51**: e324–36.

7	Sudlow C, Gallacher J, Allen N, *et al.* UK Biobank: An Open Access Resource for Identifying the Causes of a Wide Range of Complex Diseases of Middle and Old Age. *PLoS Med* 2015; **12**: e1001779.

8	Bycroft C, Freeman C, Petkova D, *et al.* The UK Biobank resource with deep phenotyping and genomic data. *Nature* 2018; **562**: 203–9.

9	Casanova F, Tian Q, Atkins JL, *et al.* Iron and risk of dementia: Mendelian randomisation analysis in UK Biobank. *J Med Genet* 2024; : jmg-2023-109295.

10	Fairley S, Lowy-Gallego E, Perry E, Flicek P. The International Genome Sample Resource (IGSR) collection of open human genomic variation resources. *Nucleic Acids Res* 2020; **48**: D941–7.

11	Mbatchou J, Barnard L, Backman J, *et al.* Computationally efficient whole-genome regression for quantitative and binary traits. *Nat Genet* 2021; **53**: 1097–103.

12	Kurki MI, Karjalainen J, Palta P, *et al.* FinnGen provides genetic insights from a well-phenotyped isolated population. *Nature* 2023; **613**: 508–18.

13	Sollis E, Mosaku A, Abid A, *et al.* The NHGRI-EBI GWAS Catalog: knowledgebase and deposition resource. *Nucleic Acids Res* 2023; **51**: D977–85.

14	Otowa T, Hek K, Lee M, *et al.* Meta-analysis of genome-wide association studies of anxiety disorders. *Mol Psychiatry* 2016; **21**: 1391–9.

15	Olafsdottir TA, Theodors F, Bjarnadottir K, *et al.* Eighty-eight variants highlight the role of T cell regulation and airway remodeling in asthma pathogenesis. *Nat Commun* 2020; **11**. DOI:10.1038/S41467-019-14144-8.

16	Roselli C, Chaffin MD, Weng LC, *et al.* Multi-ethnic genome-wide association study for atrial fibrillation. *Nat Genet* 2018; **50**: 1225–33.

17	Wuttke M, Li Y, Li M, *et al.* A catalog of genetic loci associated with kidney function from analyses of a million individuals. *Nat Genet* 2019; **51**: 957–72.

18	Sakornsakolpat P, Prokopenko D, Lamontagne M, *et al.* Genetic landscape of chronic obstructive pulmonary disease identifies heterogeneous cell-type and phenotype associations. *Nat Genet* 2019; **51**: 494–505.

19	Aragam KG, Jiang T, Goel A, *et al.* Discovery and systematic characterization of risk variants and genes for coronary artery disease in over a million participants. *Nat Genet* 2022; **54**: 1803–15.

20	Howard DM, Adams MJ, Clarke TK, *et al.* Genome-wide meta-analysis of depression identifies 102 independent variants and highlights the importance of the prefrontal brain regions. *Nat Neurosci* 2019; **22**: 343–52.

21	Bovijn J, Jackson L, Censin J, *et al.* GWAS Identifies Risk Locus for Erectile Dysfunction and Implicates Hypothalamic Neurobiology and Diabetes in Etiology. *Am J Hum Genet* 2019; **104**: 157–63.

22	An J, Gharahkhani P, Law MH, *et al.* Gastroesophageal reflux GWAS identifies risk loci that also associate with subsequent severe esophageal diseases. *Nat Commun* 2019; **10**. DOI:10.1038/S41467-019-11968-2.

23	Gharahkhani P, Jorgenson E, Hysi P, *et al.* Genome-wide meta-analysis identifies 127 open-angle glaucoma loci with consistent effect across ancestries. *Nat Commun* 2021; **12**. DOI:10.1038/S41467-020-20851-4.

24	Tin A, Marten J, Halperin Kuhns VL, *et al.* Target genes, variants, tissues and transcriptional pathways influencing human serum urate levels. *Nat Genet* 2019; **51**: 1459–74.

25	Praveen K, Dobbyn L, Gurski L, *et al.* Population-scale analysis of common and rare genetic variation associated with hearing loss in adults. *Commun Biol* 2022; **5**: 540.

26	Shah S, Henry A, Roselli C, *et al.* Genome-wide association and Mendelian randomisation analysis provide insights into the pathogenesis of heart failure. *Nat Commun* 2020; **11**. DOI:10.1038/S41467-019-13690-5.

27	Teumer A, Chaker L, Groeneweg S, *et al.* Genome-wide analyses identify a role for SLC17A4 and AADAT in thyroid hormone regulation. *Nat Commun* 2018; **9**. DOI:10.1038/S41467-018-06356-1.

28	Eijsbouts C, Zheng T, Kennedy NA, *et al.* Genome-wide analysis of 53,400 people with irritable bowel syndrome highlights shared genetic pathways with mood and anxiety disorders. *Nat Genet* 2021; **53**: 1543–52.

29	Gormley P, Anttila V, Winsvold BS, *et al.* Meta-analysis of 375,000 individuals identifies 38 susceptibility loci for migraine. *Nat Genet* 2016; **48**: 856–66.

30	Boer CG, Hatzikotoulas K, Southam L, *et al.* Deciphering osteoarthritis genetics across 826,690 individuals from 9 populations. *Cell* 2021; **184**: 4784-4818.e17.

31	Zhang H, Ahearn TU, Lecarpentier J, *et al.* Genome-wide association study identifies 32 novel breast cancer susceptibility loci from overall and subtype-specific analyses. *Nat Genet* 2020; **52**: 572–81.

32	Saevarsdottir S, Stefansdottir L, Sulem P, *et al.* Multiomics analysis of rheumatoid arthritis yields sequence variants that have large effects on risk of the seropositive subset. *Ann Rheum Dis* 2022; **81**: 1085–95.

33	Trubetskoy V, Pardiñas AF, Qi T, *et al.* Mapping genomic loci implicates genes and synaptic biology in schizophrenia. *Nature* 2022; **604**: 502–8.

34	Mahajan A, Spracklen CN, Zhang W, *et al.* Multi-ancestry genetic study of type 2 diabetes highlights the power of diverse populations for discovery and translation. *Nat Genet* 2022; **54**: 560–72.

35	De Lange KM, Moutsianas L, Lee JC, *et al.* Genome-wide association study implicates immune activation of multiple integrin genes in inflammatory bowel disease. *Nat Genet* 2017; **49**: 256–61.

36	Denaxas S, Gonzalez-Izquierdo A, Direk K, *et al.* UK phenomics platform for developing and validating electronic health record phenotypes: CALIBER. *Journal of the American Medical Informatics Association* 2019; **26**: 1545–59.

37	Calderón-Larrañaga A, Vetrano DL, Onder G, *et al.* Assessing and Measuring Chronic Multimorbidity in the Older Population: A Proposal for Its Operationalization. *J Gerontol A Biol Sci Med Sci* 2016; : glw233.

38	Yang J, Zeng J, Goddard ME, Wray NR, Visscher PM. Concepts, estimation and interpretation of SNP-based heritability. *Nature Genetics 2017 49:9* 2017; **49**: 1304–10.

39	Bulik-Sullivan BK, Loh P-R, Finucane HK, *et al.* LD Score regression distinguishes confounding from polygenicity in genome-wide association studies. *Nat Genet* 2015; **47**: 291–5.

40	Bulik-Sullivan B, Finucane HK, Anttila V, *et al.* An atlas of genetic correlations across human diseases and traits. *Nat Genet* 2015; **47**: 1236–41.

41	Mägi R, Morris AP. GWAMA: software for genome-wide association meta-analysis. *BMC Bioinformatics* 2010; **11**: 288.
